# Segmentation of the Human Tongue Musculature Using MRI: Field Guide and Validation in Motor Neuron Disease

**DOI:** 10.1101/2024.12.12.24318964

**Authors:** Thomas B Shaw, Fernanda L Ribeiro, Xiangyun Zhu, Patrick Aiken, Saskia Bollmann, Steffen Bollmann, Jeryn Chang, Kali Chidley, Harriet Dempsey-Jones, Zeinab Eftekhari, Jennifer Gillespie, Robert D Henderson, Matthew C Kiernan, Ira Ktena, Pamela A McCombe, Shyuan T Ngo, Shana T Taubert, Brooke-Mai Whelan, Xincheng Ye, Frederik J Steyn, Sicong Tu, Markus Barth

## Abstract

The human tongue is a multifunctional organ with crucial roles in speech and swallowing. However, due to its anatomical characteristics, it is difficult to measure constituent muscle volumes, and few studies have investigated this aspect thoroughly. The present study introduces a novel method and datasets for automatically segmenting the musculature of the human tongue using non-invasive T1-weighted and T2-weighted MRI. We generated accurate and robust volume estimation of five intrinsic tongue muscles and developed and validated a comprehensive volume estimation method combining AI-assisted (deep learning) segmentation and atlas fusion approaches. We showcase its clinical potential in over 200 Magnetic Resonance (MR) scans including a clinically phenotyped cohort of neurodegenerative patients with motor neuron disease (MND). We describe detailed guidelines, publicly available multi-site datasets, and an externally validated normalisation strategy that removes confounding demographic characteristics and will allow generalised application across large publicly available MRI neuroimaging datasets. Results show expected differences in people living with MND; notably, those with clinical evidence of speech and swallowing dysfunction have smaller muscle volumes. As the tongue is generally covered in neuroimaging protocols, we envision that our proposed and validated analysis pipeline will allow the post-hoc analysis of a vast number of datasets. We expect this novel work to enable the investigation of tongue muscle volume as a marker in a wide range of diseases that implicate tongue function, including neurodegenerative diseases, pathological speech disorders and cancer.

## Introduction

The tongue is a remarkable organ. Its hydrostatic muscular properties enable compression and compensatory expansion across dimensions, facilitating mastication, bolus propulsion for swallowing, the oral shaping of speech sounds [1], [2], [3], [4], and maintaining upper airway patency for breathing [5], [6]. In motor-related neurodegenerative disorders (e.g., Motor Neuron Disease [MND]/ Amyotrophic Lateral Sclerosis [ALS] [9], [11], [12], [13], [14] Parkinson’s disease, MND, Kennedy disease, multiple sclerosis, frontotemporal dementia) and following neurological injury (e.g., cerebrovascular accident, traumatic brain injury), lingual function may be impacted [15], [16], [17], [18]. Functionally, these symptoms can adversely affect speech and swallowing, with dysarthria (speech difficulties) and dysphagia (swallowing difficulties) being common clinical features [19], [20]. Clinically, tongue *structure* is visually examined for atrophy and fasciculation. As such, imaging-based markers of tongue structure would provide critical insights into the neuromuscular substrates related to lingual dysfunction. However, accurate and usable methodologies for labelling borders (segmenting) the musculature of the tongue for volumetric studies are lacking. Such methodologies would have far-reaching applications in neurodegenerative disorders (e.g., across the spectrum of MNDs [9]) to aid in the timely identification of impaired tongue musculature, informing diagnosis, prognosis, and treatment planning.

The muscles of the tongue can be categorised anatomically into interconnected groups [2], [3], [21]. The *intrinsic* tongue muscles originate and insert within the tongue. There are four paired intrinsic muscles, namely: 1) superior longitudinal, 2) inferior longitudinal, 3) transverse, and 4) vertical muscles. The *extrinsic* muscles originate from structures outside the tongue and include the 5) genioglossus, 6) hyoglossus, 7) styloglossus, and 8) palatoglossus muscles. In our present study, we focus on the superior longitudinal, the transverse, the vertical, the genioglossus, and the inferior longitudinal muscles, as the remaining muscles are too small/diverse to be characterised in conventional MR scans. These muscles act synergistically to retract, shorten, elongate, flatten, and subtly shape the tongue for speech and swallowing[22]. Despite the critical function of these muscles in, for example, speech formation, medical image segmentation and volumetric studies of the tongue musculature are challenging due to the heterogeneity of the intrinsic and extrinsic muscles, motion during scanning, and the unclear definitions between muscle boundaries and interdigitations [23]. Previous work has focused on segmentation of the entire tongue using Magnetic Resonance (MR), CT, and Ultrasound [24], [25], [26], [27], [28], with studies focused on dynamic/CINE MRI [29], [30], [31], and contour-based approaches [32]. One report [33] used deep learning-based 2D (single slice) dynamic segmentation for the vocal tract and articulators (including the whole tongue), but did not extend to individual musculature, limiting its use in clinical contexts. Another report [34] generated an atlas of the various muscles of the tongue, though to our knowledge, detailed segmentation methodology has not been released publicly.

Moreover, the tongue, like many biological structures, varies with both biological sex and age, as well as disease [35], [36] – meaning there is a need for volume normalisation strategies to ensure precise measurements between participants to examine differences in cross-sectional studies. Normalising individual tongue muscles by total tongue volume can provide relative size comparisons; but using an overall normalisation metric—such as normalising by the intra-oral cavity volume—will better account for demographic factors like height, weight, and sex by providing a standardised anatomical reference between participants. This approach is particularly important in conditions like MND, where pathology-driven changes may otherwise skew results, enabling more reliable comparisons by controlling for individual variations in oral cavity size. An overall normalisation metric (akin to Intra-Cranial Volume in neuroimaging contexts) would be beneficial for future tongue segmentation studies.

In summary, there is a clear need for a guideline for the segmentation of the human tongue (manually and/or automatically) and its musculature using MRI and an appropriate normalisation scheme to ensure the validity of cross-sectional studies of tongue volumetry. We introduce and approach each of these needs using widely used multi-contrast (T1w and T2w) MR images of the head and neck and introduce and validate a novel normalisation strategy for the musculature of the tongue. We introduce these methods and validation experiments on real-world non-neurodegenerative controls (NCs) and clinical motor neurodegenerative patients (people living with MND: plMND), demonstrating the robustness of the techniques in both populations.

## Results

### Summary of methods

We analysed MRI scans from three datasets collected between 2018 and 2023, including non-neurodegenerative controls (NCs) and people living with motor neuron disease (plMND) subtypes: Amyotrophic Lateral Sclerosis (ALS), Primary Lateral Sclerosis (PLS), and Progressive Muscular Atrophy (PMA). First, we measured volumes of five key tongue muscles—namely the superior longitudinal, the transverse and the vertical muscles combined, the genioglossus, and the inferior longitudinal—identifiable on routine MRI scans using a semi-automated AI-based segmentation protocol. We iteratively trained models using our semi-automatic segmentation method. Moreover, we introduced a novel normalisation method using intra-oral cavity (IOC) volume to account for individual anatomical differences and assessed its validity. Next, statistical analyses were conducted to explore relationships between tongue volumes and demographic variables (sex, weight, height). Finally, we performed a clinical validation of the proposed methods using clinical measures of bulbar involvement, the ALS Functional Rating Scale-Revised (ALSFRS-R), between non-neurodegenerative controls (NCs) and people living with MND (plMND). Non-parametric tests were used due to violations of parametric tests (heteroscedacity and normality).

### Tongue segmentation

The tongue volumes acquired from the semi-automatic segmentation are shown in Figure 1 and Appendix One. Following our automatic and manual segmentation method (Fig. 1), we segmented all five tongue muscles. Segmentation examples and trends across datasets are shown in Figure 1.

**Figure 1.**
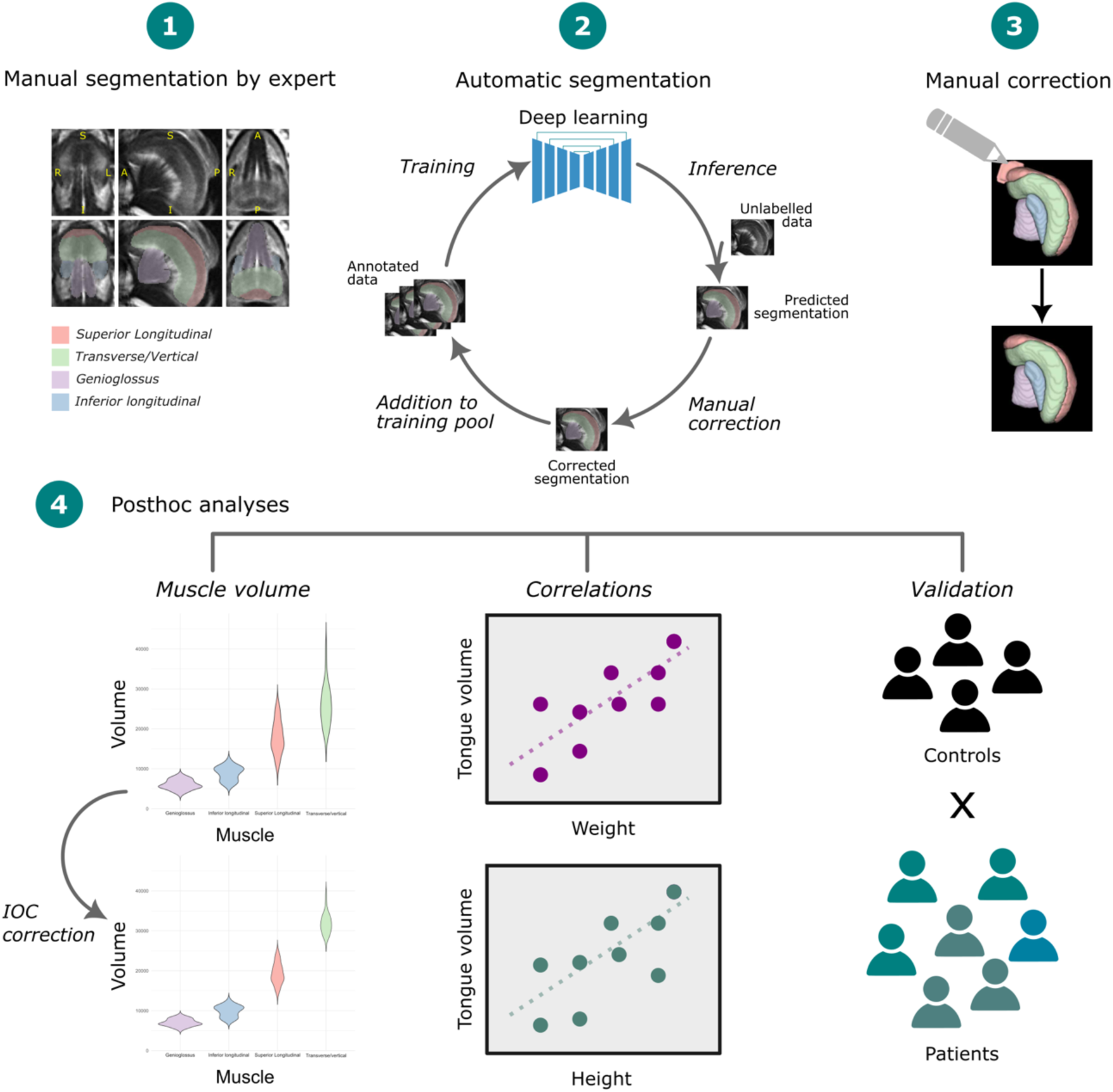
Tongue segmentation pipeline and overview. 1) Manual segmentations were created by trained staff, including five of the major tongue muscles. 2) During the labelling phase, data were iteratively added to a training pool using a variety of methods, resulting in segmentations that were quality-controlled against our manual segmentation guide and quality assurance (QA) criteria. 3) Small errors in segmentation were corrected using manual segmentation and added back to the training pool. 4) Statistical analyses for segmentation validation, intra-oral cavity (IOC) normalisation, and clinical validation in MND were conducted.

We assessed the image quality of the input images and segmentations as described in Appendix Two Table One and removed data with average scores below 2 (n=14). To ensure manual correction reliability, we conducted intra and inter-rater reliability experiments, detailed in Appendix 2, Section 3: Tables 3 and 4.

We evaluated the consistency of tongue muscle volumes across the three datasets using ANOVA, followed by Tukey’s Honest Significant Difference (Tukey HSD) post-hoc tests. The ANOVA revealed significant differences across the volumes of different muscles (F(3, 456) = 1245.483, *p* < 0.0001), between the datasets (F(2, 456) = 6.357, *p* = 0.00189), and in the interaction between different muscles and datasets (F(6, 456) = 23.861, *p* < 0.0001). These results indicated that muscle volumes significantly differ within datasets (as expected). The extent of these differences varies across datasets, suggesting population characteristics. We followed these inconsistencies up in Experiment 3 (below), where we take a normalisation factor into account.

For our normalisation experiments, we established a clinical cut-off score for bulbar involvement, to exclude participants with possible tongue atrophy from our validation experiments (see methods). We expected tongue volume between the control group and the “intact” bulbar group would be consistent and removed patients who had lost two or more points in the “Speech” or “Swallowing” ALSFRS-R bulbar sub scores. We conducted Welch Two-Sample t-tests between the groups. For each muscle, no significant differences in volumes between NCs and non-bulbar involvement plMND were found. For total volume (mean difference = −0.008, 95% CI = [−4.53, 2.99], p = 0.69), inferior longitudinal (mean difference = 0.46, 95% CI = [−0.25, 1.17], p = 0.20), superior longitudinal (mean difference = −0.49, 95% CI = [−2.02, 1.04], p = 0.53), transverse/vertical (mean difference = −0.67, 95% CI = [−2.51, 1.17], p = 0.47), and genioglossus (mean difference = −0.07, 95% CI = [−0.66, 0.53], p = 0.83).

### Intra-Oral Cavity (IOC) measurements and normalisation

We estimated the IOC to approximate head size for normalisation of demographic factors that would influence group-level statistics. Figure 2a shows our proposed normalisation approach based on estimating the intra-oral cavity. This involves measuring seven aspects of the IOC manually to estimate two elliptical frustums and calculating the resulting volumes (see Methods and Appendix 2, Section One for more detail). Normalised tongue muscle volume is determined by dividing the muscle volume by the IOC and is important to consider for removing confounding demographic variables that can skew tongue volume measurements (e.g., men having larger tongues in general).

**Figure 2.**
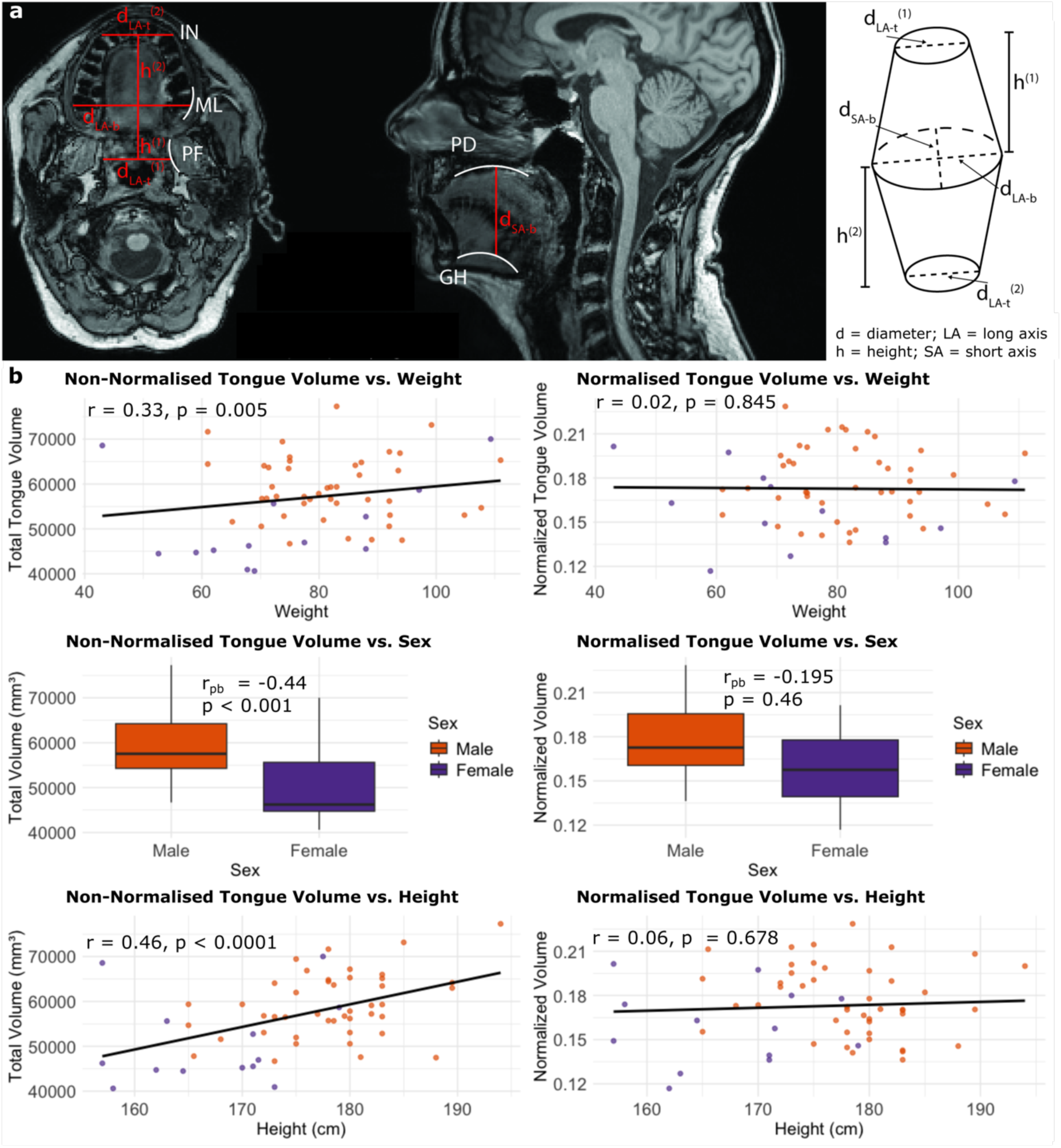
a) We measured the volumes of two frustums on each dataset to calculate the approximate Intra-Oral Cavity (IOC; i.e., the mouth) of each participant. The landmarks used to demarcate each measurement are shown in white. In red are the measurements taken, where *d* = diameter, *h* = height, *LA* = long axis, *SA* = short axis, *t* = top of frustum, *b* = bottom of frustum. In white are anatomical landmarks including *IN* = incisors, *ML* = Midline, *PF* = parapharageal fat, *PD* = Palatum Durum, *GH* = Geniohyoid. b) Effect of normalisation on total tongue volumes. (left) non-normalised tongue volumes vs. (right) normalised volumes for weight (top), sex (middle), and height (bottom), showing that without normalisation, there are significant correlations with each.

We examined the validity of our IOC methodology by first conducting Pearson’s correlation between tongue volume and IOC volume. We found a significant correlation (r = .29, p < 0.001, df = 115), suggesting that IOC and tongue volume are related in our datasets. We examined the associations between sex, weight, and height with non-normalised and IOC-normalised total tongue volume (Figure 2). Sex was significantly correlated with total tongue volume (point-biserial correlation, where Male =0, Female =1,: r_pb_ = −0.44, p < 0.001). However, no significant point-biserial correlation was observed between sex and normalised tongue volume (**Figure** 2b). A positive correlation was observed between weight and non-normalised tongue volume (r = 0.33, p = 0.005), but not with normalised volume (Figure 2b). Similarly, height was significantly correlated with non-normalised tongue volume (r = 0.46, p < 0.0001), but not after normalisation (Figure 2b).

When considering individual tongue volumes with covariates, sex (**Figure** 3) was significantly correlated with all non-normalised tongue volumes, and not with normalised tongue volumes. Due to incomplete weight and height data in datasets, correlations of these with individual tongue volumes were not conducted as reduced the sample size and statistical power would introduce bias and unreliable estimates. For completeness, to understand which demographic variables were contributing to non-normalised tongue volume, a series of regression models were run, and are expanded upon in Appendix Two Section Four.

**Figure 3.**
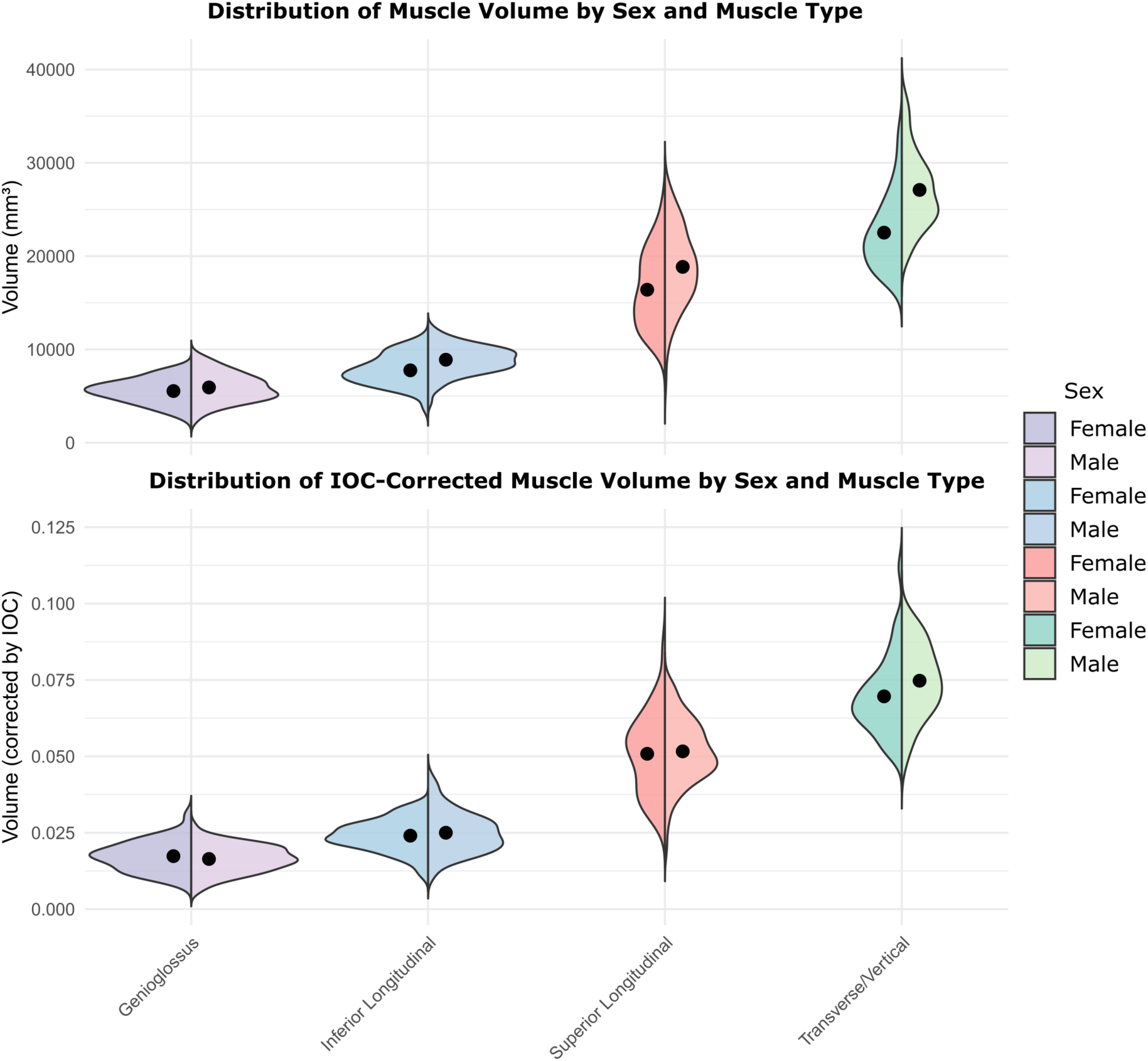
Distribution of muscle volume (top) and IOC-corrected (bottom) muscle volume by sex (female left, male right) across the five muscles. The IOC correction successfully removes the sex confounder.

### IOC correction vs relative ratio

We also correlated each IOC normalised muscle volume to the ratio of the non-normalised muscle volume to the whole tongue volume. The Superior Longitudinal muscle exhibited a strong positive correlation (r = 0.66, *p* < 0.001). Similarly, the Genioglossus showed a very strong positive correlation (r = 0.83, *p* < 0.001), the Inferior Longitudinal muscle also demonstrated a strong positive correlation (r = 0.77, *p* < 0.001), and for the Transverse/Vertical muscle, a moderate positive correlation was observed (r = 0.41, *p* < 0.001). These findings indicate a significant relationship between IOC-corrected and ratio volumes across participants, suggesting that either normalisation technique may be used for volume estimation, depending on the research question (e.g., in longitudinal analyses, the relative loss/gain of muscle may be more relevant than absolute).

### Tongue position experiments

We noticed that some participants had vastly different tongue postures in their MR scans, potentially influencing the estimation of segmentation volumes. We noticed the tongue rests in the IOC in a range of positions or postures, depending on anatomical features, muscular heterogeneity, and personal preferences. To investigate potential changes to tongue and muscle volumes due to tongue positioning, we conducted a postural experiment on one control with five tongue positions (see Figure 4 and full details in Appendix 2, Section 5).

**Figure 4.**
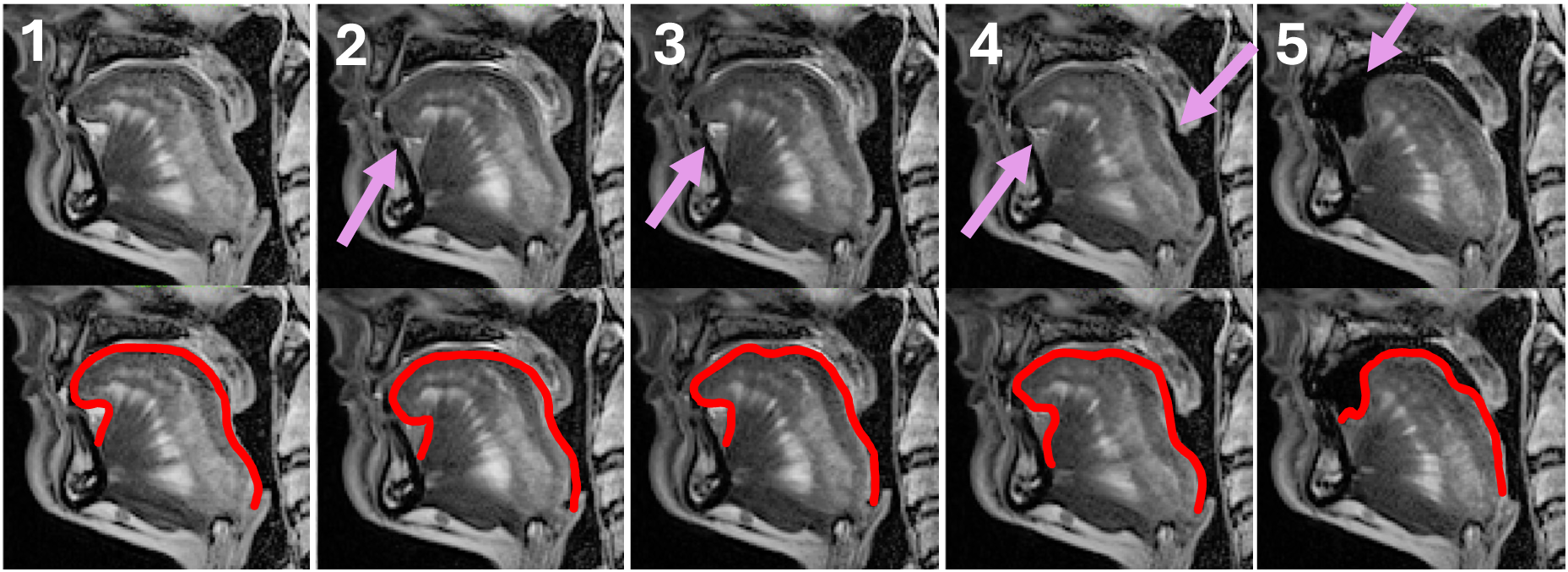
Sagittal sections of the five tongue movement positions. (Top) Pink arrows demarcate areas of changed posture; red outlines show the border of the superior longitudinal muscle.

The average percentage changes in muscle volumes between positions and the mean percentage change from each position to the subsequent one are shown in Table 1, and were <2.5%. A Repeated Measures ANOVA revealed no significant difference in muscle volumes across any of the five positions (*F* (4, 12) = 0.6804, *p* = 0.6187). This suggests that the changes observed in muscle volumes are not statistically significant between tongue positions.

**Table 1.** Muscle volumes of one control participant measured five times with varying tongue positions. The average change (measured between each of the 5 positions) was less than 2.5%, indicating the position of the tongue in the mouth does not affect our segmentation results significantly.

| *Muscle Labels* | *Position one* | *Position two* | *Position three* | *Position four* | *Position five* | *Average change %* |
| --- | --- | --- | --- | --- | --- | --- |
| Superior Longitudinal | 18631.7 | 18321.9 | 18474 | 18410 | 18885.6 | 0.35% |
| Transverse/Vertical | 11732 | 12370.9 | 12999.2 | 14458.4 | 12683.8 | 2.37% |
| Inferior Longitudinal | 4505.09 | 3431.94 | 3720.7 | 3777.02 | 4025.34 | 1.83% |
| Genioglossus | 5533.18 | 5081.09 | 5310.46 | 5272.06 | 5184 | 1.51% |

### Clinical relevance in ALS

The spectrum of MNDs can be classified into different diagnoses, with different phenotypic expression. Using IOC-normalised data, we conducted a PERMANOVA analysis (non-parametric MANOVA [37]), which revealed a significant effect of diagnosis on tongue muscle volumes (R² = 0.100, F(3,161) = 5.981, p < 0.001) suggesting that the multivariate combination of muscle volumes (i.e., the simultaneous combination of muscles, rather than any individual muscle change) differ significantly across diagnostic groups considered as part of the MND spectrum (see **Figure 5a**). Subsequently, pairwise PERMANOVA between the combined plMND group (ALS, PLS, Flail Limb/PMA, combined) and NCs revealed a significant difference (F = 11.69, R² = 0.067, p < 0.001, Bonferroni-adjusted p = 0.001), indicating that muscle volume patterns were significantly different between plMND and NCs. Further pairwise PERMANOVA comparisons between individual diagnostic groups showed the following: Control vs ALS: Significant differences (F = 12.46, p < 0.001, Bonferroni-adjusted p = 0.006). PLS vs ALS: Significant differences (F = 4.82, p = 0.007, Bonferroni-adjusted p = 0.042). Control vs PLS, Control vs Flail Limb/PMA, and Flail Limb/PMA vs ALS: No significant differences were detected after Bonferroni correction (p > 0.05).

**Figure 5.**
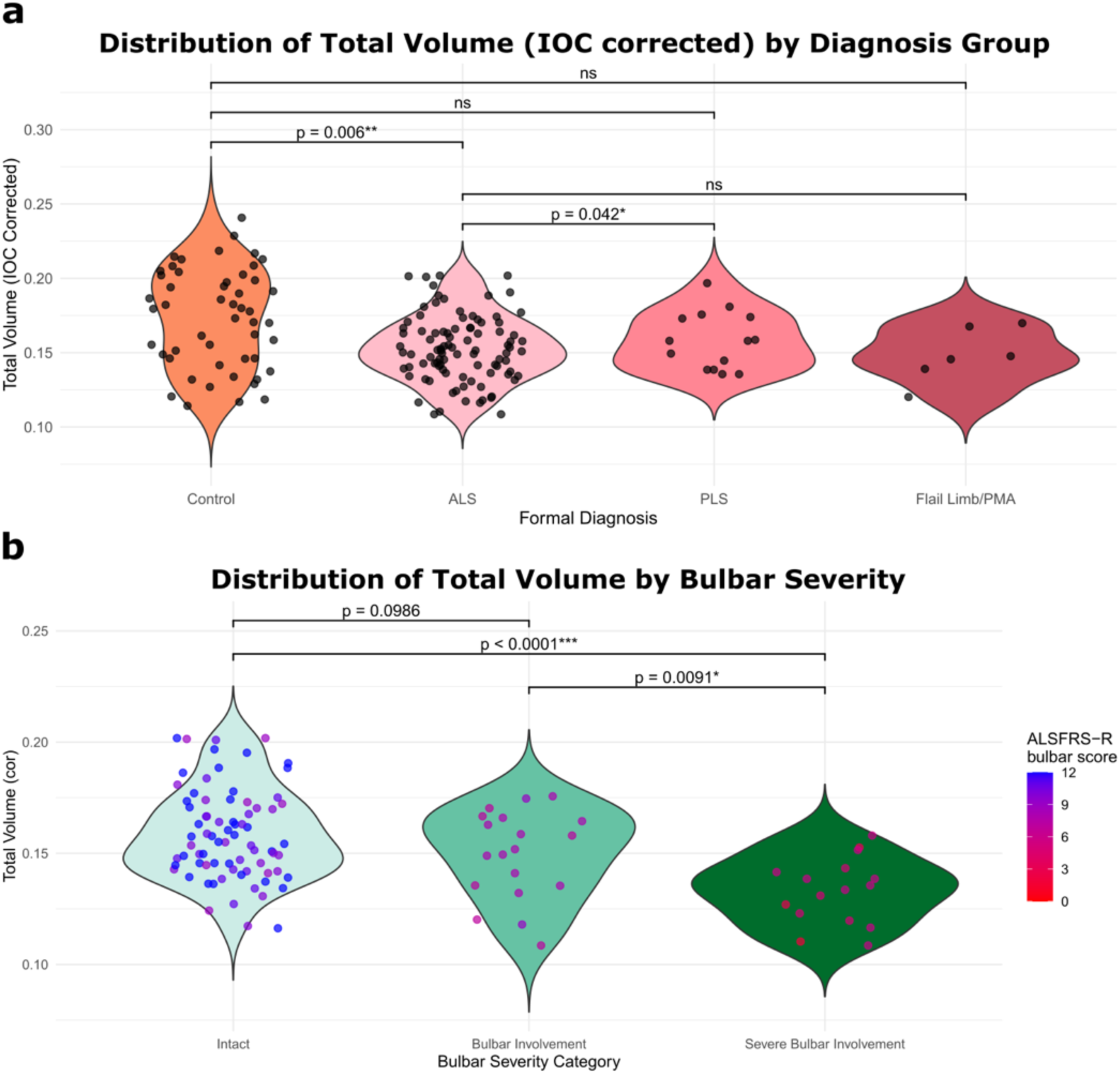
a) Distribution of IOC-corrected total volumes by the different diagnostic groups including Amyotrophic Lateral Sclerosis (ALS), Primary Lateral Sclerosis (PLS), and Flail limb phenotype/ Progressive Muscular Atrophy (PMA). Significant differences in the 2-way PERMANOVA tests are indicated: N.B. multivariate (multiple constituent tongue volume) trends may be hidden within the data. b) Shows differences between patients with clinically defined bulbar/severe bulbar involvement and those without (“Intact”). There was a marginally non-significant (p = 0.0986) difference between “Intact” and “Bulbar involvement”. All other comparisons were significant.

#### Bulbar involvement

A one-way ANOVA indicated a significant effect of bulbar severity on total tongue volume (F(2, 109) = 11.03, p < 0.001). Bonferroni-corrected pairwise t-tests revealed that plMND with “Severe bulbar involvement” had significantly lower total tongue volumes compared to those with no clinical signs of bulbar involvement “Intact” (p < 0.001) and those with “Bulbar involvement” (p < 0.01; Figure 5b). No significant difference was found between the “Intact” and “Bulbar involvement” groups (p = 0.0986). These results suggest that total tongue volume is most reduced in plMND with severe bulbar involvement.

## Discussion

We present a novel segmentation and normalisation method for the tongue *in vivo* using standard T1w or T2w 3T MR images. We have rigorously evaluated and validated our methods and segmentations with respect to both their biological variability and segmentation reliability and validated these in a clinical context. Our methods and particularly our atlases[38] are of value to the medical imaging community, as they provide ground truth labels for future studies. We first provide our general framework for tongue segmentation, including a guideline for the manual segmentation of the tongue and its musculature using MRI (Appendix 1) and publicly available datasets and atlases (available at https://osf.io/wt9fc/). We validated our method rigorously by examining intra/inter rater reliability experiments and conducting a tongue position experiment, to replicate real-world scenarios of tongue position alterations. We provide an appropriate normalisation scheme to ensure validity of cross-sectional studies of tongue volumetry in the future and have validated our segmentation methods in a clinical context (MND), where bulbar dysfunction and tongue muscle degeneration often occur.

We were able to reliably segment five large muscles within the tongue routinely with this method. In the case of the transverse and vertical muscle, the segmentation was combined due to the tightly woven muscle fibres. However, other smaller muscles posed challenges for yielding reliable segmentations, either manually or in an automated fashion. For example, the hyoglossus was segmented in our protocol, but consistently poor segmentations on scans necessitated its removal in our results. Future works could examine the intensity and shape-based characteristics of these muscles in training data, which can inform segmentation models with appropriate priors.

Our segmentation method shows promise for this type of analysis, where the vast heterogeneity of biological tissue demands a heterogeneous approach. We iteratively trained models using our semi-automatic segmentation method, progressively refining the training set with manual corrections. This approach significantly reduces the manual effort required and improves model performance over time[38]. Unlike previous studies that have not released their data and detailed segmentation methodologies [29], we provide comprehensive guidelines for manual segmentation in Appendix 1 and leverage AI-based segmentation tools to enhance efficiency and accuracy.

Specifically, our segmentation approach integrates MONAI Label and Joint Label Fusion (JLF), a common atlas-based segmentation algorithm[39]. MONAI Label[40], integrated within 3D Slicer[41]. By combining MONAI and JLF, our method benefits from the robustness of deep learning models and the generalisability of atlas-based segmentations for edge cases. By making our data and methodologies publicly available, we provide a valuable resource for the research community.

### IOC measurements and normalisation

IOC normalisation experiments provide insights into relationships between tongue volume and demographic variables. The initial correlation analyses revealed significant associations between sex and non-normalised tongue volume, and between weight and non-normalised tongue volume. However, these correlations were no longer significant after normalising the tongue volume by the IOC. Previous reports have shown strong correlations with sex, height, and weight with tongue volume [36] and indeed our own dataset reflected these trends. Introducing our normalisation strategy mitigates the effects of biological covariates, strengthening our conclusions. Future work may attempt to automatically segment the IOC (e.g., using AI-based segmentation), removing the need for manual measurements.

### IOC correction and Ratio correction

We correlated each IOC-normalised muscle volume to the ratio of the non-normalised muscle volume to the whole tongue volume and found strong correlations. Our findings indicate that either method may be used depending on the research question. For example, examining longitudinal change in muscle volume may be better examined using the ratio method, rather than an IOC correction. By accounting for individual differences in head size, the IOC normalisation process enhances the accuracy of volumetric analyses of tongue musculature, especially in a cross-sectional context.

#### Tongue Position Experiment

The average percentage changes in muscle volumes between consecutive positions indicated minimal variation. We found that, while keeping the segmentation method consistent, variations observed in muscle volumes due to different tongue positions are negligible. While this may mitigate some of the longitudinal variation in tongue segmentation studies, uniquely longitudinal segmentation methods will be required in future works [42], [43].

### Clinical relevance

This study has important clinical implications, particularly for neurodegenerative/neuromuscular diseases such as MND/ALS, where bulbar involvement predicts earlier death [44], [45]. Accurate volume measurement of tongue musculature may assist in the early identification of bulbar dysfunction, which is critical for timely diagnosis and intervention. This may improve prognostic evaluations and help tailor interventions more effectively.

In this study, we have shown gross volumetric evidence in a retrospective cohort for tongue degeneration in MND and MND subtypes. Results showed expected changes with tongue musculature that indicated degeneration plMND, and especially with those who had decreased clinical scores that reflect tongue dysfunction. Our results are the first step towards a deeper investigation into the role of the tongue in MND/ALS and serve as a clinical validation to the segmentation methods provided here. We are aware that results shown here are by no means exhaustive, and future works will explore the relationships between tongue musculature and MND/ALS more thoroughly.

### Limitations

Although our approach leverages AI-based tools, manual corrections were used to refine some of the segmentation results (especially in the training phase of the AI-model and building of atlases). This reliance on manual input can be time-consuming and introduces potential variability due to differences in rater expertise and interpretation. However, we have provided an extensive guide for segmentation in Appendix 1, and hope that our dataset release [38] will enable e.g., transfer learning or a stepping-off point for future work.

Our current method reliably segmented five large tongue muscles, but smaller muscles could not be segmented with high confidence. This is particularly limiting for work interested in e.g., the vagus nerve (CNX) innervated palatoglossus. This limitation may hinder comprehensive analysis of tongue musculature, particularly in conditions where smaller muscles play a role.

### Conclusion

This study presents a detailed methodology for segmenting and normalising the human tongue musculature using MRI across a diverse dataset of over 200 MR scans, including a clinically characterised cohort of neurodegenerative patients with MND. Given that the tongue is frequently captured in neuroimaging protocols, our analysis pipeline enables retrospective analysis across myriad datasets. By offering comprehensive guidelines, openly accessible multi-site datasets, and an externally validated normalisation strategy, this work facilitates broader application across neuroimaging datasets. The method holds significant clinical potential for early detection and monitoring of neurodegenerative diseases such as MND/ALS, with our results indicating significant differences in individuals with MND, particularly those with clinical manifestations of speech and swallowing dysfunction, supporting the method’s potential utility as a biomarker for disease monitoring. We anticipate that this work will broaden the use of tongue muscle volume as a marker for diseases impacting speech and swallowing functions, including neurodegenerative diseases, speech disorders, and cancer.

## Methods

### Data sets used

Three datasets collected between 2018-2023 were used for this study. All studies were approved by their relevant Human Research Ethics Committees, and written informed consent was obtained. All studies had a focus on MND and patients were diagnosed by experienced neurologists (RDH; MCK) according to clinical phenotype, and diagnostic criteria[46].

We included data from both controls and MND clinical phenotypes with a classical presentation of upper (UMN) and lower (LMN) motor neuron involvement (i.e., Amyotrophic Lateral Sclerosis [ALS]) and regionally limited presentations (i.e., Primary Lateral Sclerosis [PLS] – UMN only involvement; Progressive Muscular Atrophy [PMA] – LMN only involvement). Clinical validation experiments were conducted by excluding data sets from individuals with significant symptoms that affected their tongue muscles (known as “bulbar” symptoms), which was measured by a decrease in scores on the domains of the revised ALS functional rating scale (ALSFRS-R) relating to ‘bulbar dysfunction’[47]. Here, a clinical cutoff of a drop in two or more points in either the “speech” or “swallowing” subs-cores (from a possible four) was considered “bulbar involvement”, while six or more was considered “severe bulbar involvement”. We chose these cutoffs from clinical experience and in agreement with [48], [49]. To examine the validity of our clinical cut-off for bulbar dysfunction in MND, we compared volumes between non-neurodegenerative controls (NCs), and those participants who had a diagnosis, and their ALSFRS-R bulbar sub-scores did not fall into the “severe/ bulbar involvement” groups.

#### Data

Inclusion and exclusion parameters for each study are included in Figure 6. Of note are the two streams of 1) segmentation validation and 2) clinical validation within this work, which used independent imaging samples. All imaging parameters and demographics for each dataset for the segmentation validation experiments are presented in Table 2.

**Figure 6.**
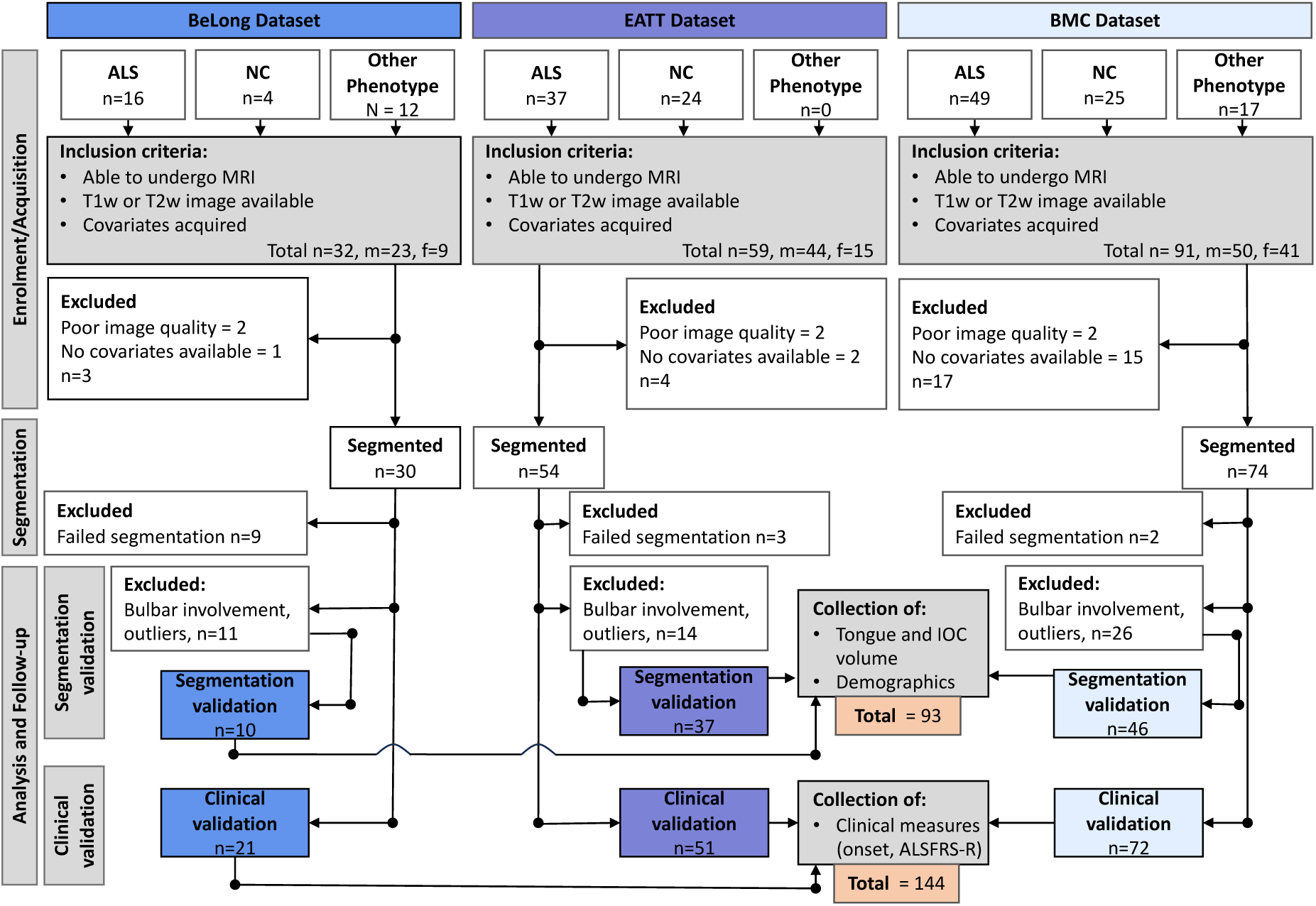
Diagram of inclusion and exclusion of participants from each of the datasets. We began with 201 independent observations. Enforcing our exclusion criteria resulted in 123 tongue segmentations for analysis in this work. NC = Non-neurodegenerative control, ALS = Amyotrophic Lateral Sclerosis, IOC = Intra-Oral Cavity. ‘Other phenotype’ includes other Motor Neuron Disease phenotypes. ‘Other phenotype’ and ALS comprise the plwMND sample. Note that our segmentation validation and clinical validation experiments are drawn from the same pool, with n=93 for the former and n=144 for the latter.

**Table 2.**
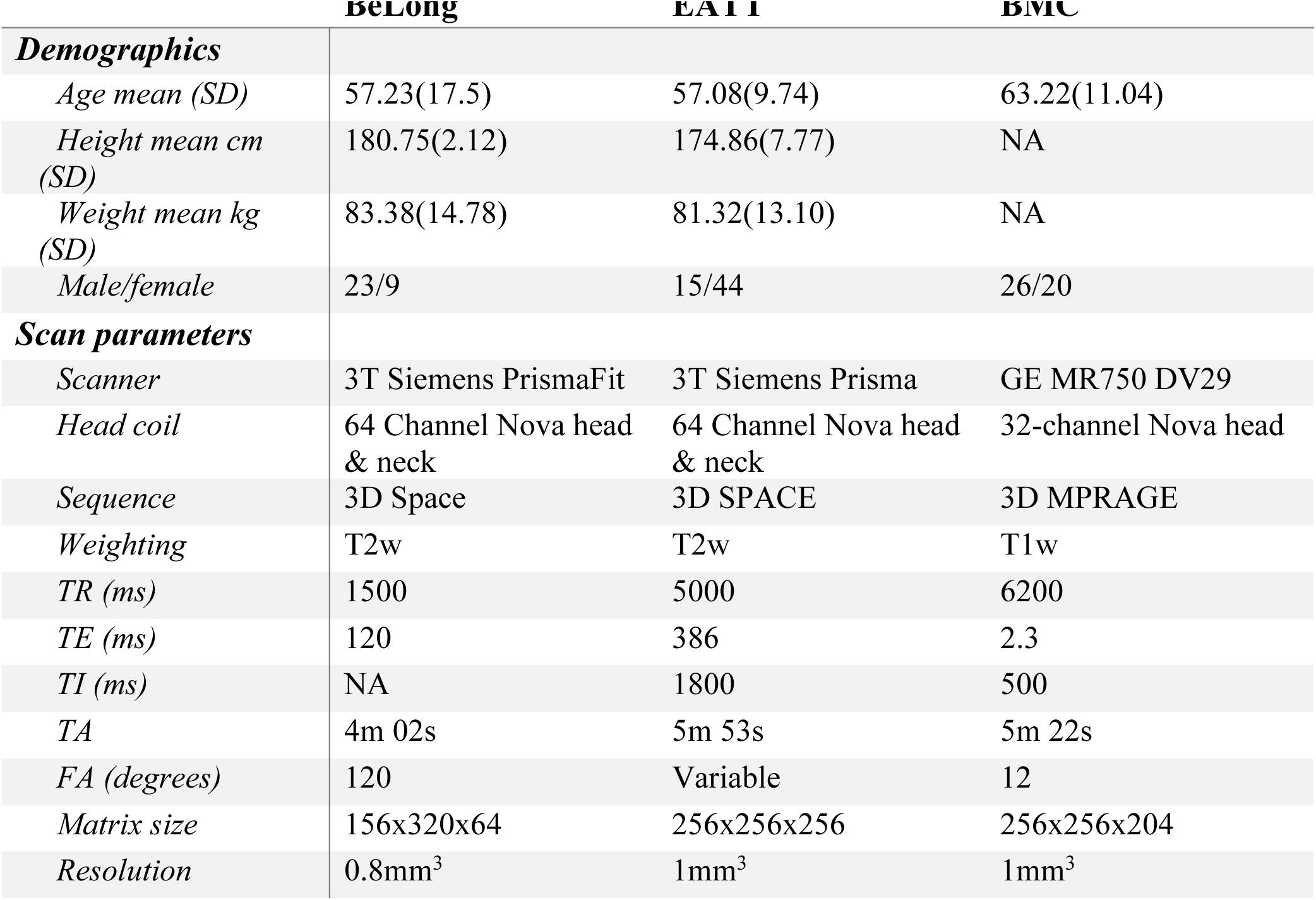
General demographic information and imaging parameters for each dataset.

Clinical/demographic metrics are described in Table 3. Data from the Biomarkers of Long surviving MND (**BeLong**) study were collected between 2020-2023 at the University of Queensland, Centre for Advanced Imaging. Details of the **EATT** MND dataset has been described previously [50] and was collected at Herston Imaging Research. Finally, the Sydney Brain and Mind Centre (**BMC**) MND dataset was collected at the BMC at The University of Sydney, Australia.

**Table 3.** Demographic information and mean relevant volumes for each clinical subgroup across datasets.

|  | *BeLong* |  |  | *EATT* |  |  | *BMC* |  |  |
| --- | --- | --- | --- | --- | --- | --- | --- | --- | --- |
|  | ALS | Control | PLS/PMA | ALS | Control | PLS/PMA | ALS | Control | PLS/PMA |
| **Demographics** |  |  |  |  |  |  |  |  |  |
| *Age years (SD)* | 59.92<br>(12.76) | 44.02<br>(26.41) | 65.26 (9.52) | 59.47 (9.6) | 54.75<br>(11.18) | 62.47 (8.98) | 62.13<br>(10.59) | 63.96<br>(9.91) | 64 (NA) |
| *Height cm (SD)* | 175.91<br>(7.27) | 180.67<br>(11.02) | 178.86 (4.14) | 173.39<br>(7.76) | 176.19<br>(7.2) | 163 (7.07) | NA | NA | NA |
| *Weight Kg (SD)* | 81.18<br>(15.51) | 77.67<br>(16.62) | 85.24 (15.6) | 83.53<br>(16.87) | 87.98<br>(18.63) | 77.23 (11.68) | NA | NA | NA |
| *Sex (Female/Male)* | 10/1 | 2/1 | 5/1 | 22/6 | 6/1 | 2/1 | 29/17 | 26/20 | 1/0 |
| **Volumes** |  |  |  |  |  |  |  |  |  |
| *Trans. vol. IOC cor. mm<sup>3</sup> (SD)* | 0.077<br>(0.01) | 0.077<br>(0.017) | 0.075 (0.012) | 0.07<br>(0.011) | 0.079<br>(0.011) | 0.064 (0.017) | 0.061<br>(0.01) | 0.073<br>(0.015) | 0.066 (NA) |
| *Trans. vol. mm<sup>3</sup> (SD)* | 28748.8<br>(5665.01) | 28326.57<br>(11400.12) | 29648.75<br>(5697.35) | 23212.14<br>(3466.93) | 25114.67<br>(3324.44) | 19440 (2217.49) | 23066.18<br>(4525.87) | 25315.33<br>(5366.9) | 24274 (NA) |
| *S. long. vol IOC cor. mm<sup>3</sup> (SD)* | 0.045<br>(0.011) | 0.045<br>(0.018) | 0.044 (0.01) | 0.047<br>(0.009) | 0.056<br>(0.01) | 0.04 (0.01) | 0.053<br>(0.008) | 0.053<br>(0.013) | 0.071 (NA) |
| *S. long. vol. mm<sup>3</sup> (SD)* | 16575.21<br>(4536.1) | 15115.61<br>(2843.7) | 16977.63<br>(2883.63) | 15628.61<br>(4273.27) | 17842.48<br>(2930.24) | 12015.5<br>(1636.95) | 19909.58<br>(3853.25) | 18575.09<br>(4776.88) | 26999 (NA) |
| *Genio. vol. IOC cor. mm<sup>3</sup> (SD)* | 0.012<br>(0.004) | 0.016<br>(0.006) | 0.014 (0.001) | 0.015<br>(0.004) | 0.018<br>(0.004) | 0.019 (0.001) | 0.016<br>(0.005) | 0.018<br>(0.005) | 0.02 (NA) |
| *Genio. vol. mm<sup>3</sup> (SD)* | 4621.17<br>(1588.72) | 5325.31<br>(952.67) | 5499.61<br>(1046.95) | 5087.89<br>(1194) | 5803.81<br>(957.96) | 6211.5 (2761.25) | 6051.81<br>(2161.3) | 6166.95<br>(1624.37) | 7520 (NA) |
| *I. long vol. IOC cor. mm<sup>3</sup> (SD)* | 0.019<br>(0.004) | 0.02 (0.007) | 0.02 (0.004) | 0.026<br>(0.007) | 0.03<br>(0.005) | 0.024 (0.013) | 0.02<br>(0.005) | 0.025<br>(0.005) | 0.022 (NA) |
| *I. long vol. mm<sup>3</sup> (SD)* | 6898.78<br>(1764.19) | 6635.69<br>(1066.26) | 7857.81<br>(1205.26) | 8554<br>(2198.75) | 9701.95<br>(1567.87) | 6820.5 (1139.15) | 7374.78<br>(2219.78) | 8440.68<br>(1682.63) | 8176 (NA) |
| *IOC vol mm<sup>3</sup> (SD)* | 370169.14<br>(44254.87) | 369159.64<br>(141397.98) | 396228.26<br>(56191.64) | 338895.39<br>(68349.39) | 326335.48<br>(65904.15) | 317915.51<br>(119418.7) | 381186.04<br>(73963.29) | 350915.19<br>(67095.9) | 370185.44 (NA) |
| **Clinometric details** |  |  |  |  |  |  |  |  |  |
| *ALSFRS-R total (SD)* | 40.45 (3.3) | NA | 30.43 (8.04) | 37.33 (6.1) | NA | 37 (0) | 39.26 (5.2) | NA | NA |
| *ALSFRS-R bulbar (SD)* | 10.55<br>(1.44) | NA | 9.86 (2.54) | 10.18<br>(1.83) | NA | 85 (2.12) | 9.35 (2.99) | NA | NA |
| *Bulbar onset count* | 1 | NA | 0 | 5 | NA | 1 | 13 | 0 | 0 |

#### Manual segmentation guide

All manual segmentations and manual corrections were completed in line with our manual segmentation guide (Appendix 1). This comprehensive guide to manual tongue segmentation is designed to be user-friendly for researchers, regardless of their anatomical knowledge of the tongue. The musculature of the tongue extends beyond the muscles that are annotated and described in our approach. However, robust manual identification and delineation of their boundaries (i.e., non-segmented anatomy) were considered too difficult from routine clinical whole-brain MR scans. Our approach considered real-world practicality, given the conditions upon which much MR tongue data are incidentally acquired, namely: 1) a T1w or T2w 3D MR image including the mouth and preferably the entire jaw; 2) Image quality is sufficient to view the borders of the tongue, and that the gross anatomical structures of the intrinsic musculature of the tongue are visible to some degree. The *intrinsic* tongue muscles (controlling the shape of the tongue) that were segmented were the superior longitudinal, inferior longitudinal, transverse, and vertical muscles. In our protocol, transverse and vertical were combined due to the complex interwoven fibres of the two. The *extrinsic* muscles (originating from structures outside the tongue and controlling the position of the tongue) that were segmented were the genioglossus and hyoglossus.

To ensure the accuracy and reliability of our segmentations, we developed a set of rating criteria for the quality of the tongue image. These are laid out in Appendix Two, Table One. All images were rated manually using adapted methods from [51] and [52]. We also developed a set of rating criteria for the segmentation of the tongue. We used this rating system to include and exclude segmentations from our analysis. Unless image quality is high (per our QA procedure), the hyoglossus muscle was nearly impossible to differentiate from surrounding muscle and was excluded in all our analyses.

To assess the consistency of segmentation volumes across the datasets and among different muscles, a two-way Analysis of Variance (ANOVA) was conducted to examine the main effects of dataset and muscle type on segmentation volume and their potential interaction effect.

### Semi-automated tongue segmentation

Beginning with the BeLong dataset T2w images, three tongue volumes from three scans were manually annotated by XZ (Medical Principal House Officer with 4 years medical experience). XZ was guided by consultant radiologist JG (12 years MR radiological experience). XZ subsequently corrected and labelled 6 additional scans. TBS (10 years medical imaging experience), and FLR (6 years medical imaging experience) were responsible for manual labelling and correcting downstream segmentations (∼60% of all segmentations), under the guidance of XZ.

We leveraged MONAI Label [53] to interactively annotate nine additional scans from the BeLong study[38]. Then, we used our segmentation model training implementation, in addition to these 12 labelled data, to train a segmentation model and predict tongue muscle segmentation on new unlabelled data. These initial predictions were further refined with manual correction and were leveraged with different approaches to speed up data annotation, including test-time adaptation (TTA) and Joint Label Fusion (JLF) (refer to [38] for more details). Each of these approaches for generating automatic segmentations, i.e., pre-trained model, TTA, and JLF, were used to grow the pool of annotated and manually corrected data for a new iteration of segmentation model training (Figure 1, step 2).

We iteratively bootstrapped the best segmentation out of the three methodologies to manually correct the segmentation if required. The new labelled data were then used to train a new segmentation model for the following iteration. We repeated this process until all data were segmented, and segmentation quality assured (as per our QA procedure was satisfactory or above). Our segmentation methods were intentionally multi-modal, as it was necessary to leverage the strengths of each method, given the variety of data (patient and control, different scanners, different MR contrasts).

### Validation of segmentation: Intra/inter-rater reliability

To confirm the reliability of our initial segmentation method, we implemented checks for consistency within and between raters using inter and intra-rater reliability experiments. The details for which are included in Appendix Two, Table Three and Four.

### Statistical analysis

#### Code

After extracting all tongue volumes using NumPy[54] and nibabel[55] via Python3.10[56], all downstream statistical analyses were performed using R statistical software[57]. All code is available at https://github.com/thomshaw92/TongueSegMND.

### Intra-Oral Cavity (IOC) measurements and normalisation

Tongue size and shape is heterogeneous in both healthy controls and in patients. We introduce a novel method for normalising the musculature of the tongue to the size of the individuals’ head. We developed a relatively quick (∼30s per scan) method for estimating the intra-oral cavity (IOC)[58]. This involves measuring seven aspects of the IOC manually to estimate two elliptical frustums and calculating the resulting volumes (see Fig 3) that contain most of the IOC, excluding the pharynx. Full instructions for this calculation are included in Appendix Two, Section One. Correlations between demographic variables and pre- and post-normalisation were conducted to establish the validity of the technique.

### Clinical relevance in ALS

We conducted preliminary studies into the usefulness of the tongue segmentation strategy in Motor Neuron Disease (MND) subtypes including Amyotrophic Lateral Sclerosis (ALS), Primary Lateral Sclerosis (PLS), and Progressive Muscular Atrophy (PMA). After our segmentation validation experiments, we included the remaining participants from the three datasets and conducted two clinical validity checks, namely:

#### Case-control comparisons

We assessed whether people living with MND (plMND) and the subtypes of MND (PLS, PMA, ALS) had altered tongue volume compared to those without. Due to the non-normality of the individual tongue muscle data, a multivariate permutational analysis of variance (PERMANOVA) was performed to assess the overall differences in tongue muscle volumes across diagnostic groups. The analysis included the five corrected tongue muscle volumes, while the formal diagnosis (Control, ALS, PLS, and Flail Limb/PMA) was treated as a factor. Euclidean distance was used as the distance metric, and the analysis was conducted with 999 permutations using the adonis2 function in R. Pairwise comparisons were conducted to examine differences between the combined plMND group (ALS, PLS, and Flail Limb/PMA) and NCs. These pairwise comparisons were carried out using the pairwise.adonis function with Bonferroni correction for multiple comparisons. Additionally, individual pairwise comparisons between each diagnostic group (Control, ALS, PLS, and Flail Limb/PMA) were performed to further investigate significant differences in multivariate muscle volume patterns.

#### Bulbar involvement

We compared plMND on corrected tongue volumes to determine whether bulbar involvement (as assessed using the ALSFRS-R bulbar sub-scale) was related to total corrected tongue volume. “Severe” “Bulbar involvement”, and “Intact” were defined as before. As overall corrected volume met assumptions for frequentist statistics, a one-way ANOVA was conducted with pairwise t-tests and Bonferroni corrections for follow-ups.

## Supporting information

Appendix One

Appendix Two

## Acknowledgements

TBS, JC, ST, MCK, RH, BMW, PA, PAM, STN, and FJS are supported by Motor Neurone Disease Research Australia (MNDRA). TBS, HDJ, KC, RDH, MCK, PAM, STN, FJS, ST, and MB are supported by Fight Motor Neuron Disease (FightMND). Data collection for the EATT MND data set is supported by a Wesley Medical Research grant (2-17-07) to FJS, STN and RDH. JC, KC, ZE, and XY are supported by the UQ Graduate School Scholarship (RTP). TBS, PAM, RDH, MCK, ST, MB are supported by the National Health and Medical Research Council. HDJ and MB are supported by the Australian Research Council. SB, XY and FLR acknowledge funding through an ARC Linkage grant (LP200301393). ST acknowledges Lenity Australia. The authors acknowledge the National Imaging Facility, human imaging staff at the Centre for Advanced Imaging, Herston Imaging Research Facility, and The Brain and Mind Centre. The authors thank all participants involved in this research.

## Data availability statement

All non-neurodegenerative control (NC) data and their corresponding segmentations is available in the Open Science Framework database (https://osf.io/wt9fc/). All identifying information has been removed from the dataset. We have also included a reference template and cropped tongue region for registration.

Patient data is available upon request provided appropriate HREC agreements are in place.

## Code availability statement

All code is available at https://github.com/thomshaw92/TongueSegMND including developed tools, methods for analysis, statistical analysis, and code for segmentation. A deep learning-based segmentation model will be trained on publicly available data (https://osf.io/wt9fc/) and made available upon publication of this manuscript.

