## Appendix One for "Segmentation of the Human Tongue Musculature Using MRI: Field Guide and Validation in Motor Neuron Disease"

### Overview of tongue segmentation in MRI & how-to/guide.

Xiangyun (Xavier) Zhu and Thomas B Shaw


The tongue plays a vital role in several fundamental activities, such as speech, mastication, swallowing, and taste perception. Understanding its complex muscular structure is critical in a range of clinical applications, from diagnosing pathological conditions to planning surgical interventions.

T1w images provide excellent anatomical detail, with high contrast between fat and water-containing tissues. Conversely, T2w images offer high contrast between different types of soft tissues and are particularly adept at highlighting areas of oedema, inflammation, or pathology. A multimodal (T1w + T2w) approach to manual segmentation is preferred.

We have chosen to focus on six key muscles in our segmentation process: the superior longitudinal, transverse, vertical, genioglossus, inferior longitudinal, and hyoglossus. These muscles were selected because of their larger size, distinct appearance, relative ease of identification in MRI scans, and significant contribution to tongue functionality.

In our protocol, the transverse and vertical are combined.

**Key muscles and their functions**

- **Superior Longitudinal** (yellow)
  - Helps to curl up the sides of the tongue for activities such as swallowing.
  - Assists in retracting or elongating the tongue.
  - Found on the upper surface of the tongue
- **Transverse/Vertical** (green)
  - Narrows and elongates the tongue, enabling precision movements during speech and swallowing.
  - Lies in the middle of the tongue, surrounded by other muscles
- **Genioglossus** (blue)
  - Protrudes and depresses the tongue, playing a crucial role in opening the airway.
- **Inferior Longitudinal** (salmon)
  - Retracts and helps in curling the tongue's tip.
  - Runs along the tongue's length and is more pronounced laterally.
- **Hyoglossus** (red)
  - Depresses and retracts the tongue, contributing to the formation of a clear oral cavity.
  - Lies alongside the submandibular and sublingual salivary glands and the mylohyoid muscle

In the following sections, we show the process of segmenting these muscles on T2w MRI scans, providing a step-by-step guide for each muscle group. A similar process is used for T1w MRI scans, the only difference being the contrast/signal intensity inherent to the scan.

### Other muscles (not included in this protocol) of the tongue and mouth.

Beyond the six muscles (superior longitudinal, transverse, vertical, genioglossus, inferior longitudinal, and hyoglossus) you are segmenting, other muscles are also involved, including the styloglossus, palatoglossus, and mylohyoid.
However, some muscles are challenging to distinguish and segment in MRI scans for various reasons:

- **Styloglossus**: This muscle retracts and elevates the tongue and is relatively thin compared to the other muscles. It is challenging to identify due to its size and its proximity to other muscles such as the hyoglossus.
- **Palatoglossus**: This muscle elevates the posterior part of the tongue and lowers the soft palate, forming part of the oral cavity's anterior pillar. Like the styloglossus, it's thin and nestled closely between other larger muscles, making it difficult to identify and differentiate on standard T1w/T2w.
- The **mylohyoid** muscle is a thin, flat muscle that forms the floor of the mouth and supports the tongue. While it is not technically a muscle of the tongue, it works closely with the tongue muscles and plays a crucial role in swallowing and speaking. Its anatomy is quite distinct, as it forms a sort of diaphragm for the oral cavity, stretching between the mandible (lower jawbone) and the hyoid bone. It is characterized by a broad, thin, rectangular shape and appears as a hypointense band on T2w MRI, located in the floor of the mouth below the tongue.

The small size and proximity of these muscles make distinguishing them from each other and the surrounding tissues difficult. Further, the inherent intensity characteristics of these muscles on T1w and T2w images are not markedly different from each other or the surrounding tissues.

### 1) Superior longitudinal (labelled yellow)

#### General overview

- The superior longitudinal muscle forms the upper part of the tongue. On T2w MRI, this muscle is seen as a hypointense band spanning the superior aspect of the tongue, running anteroposterior (from front to back). Key landmarks for identifying the superior longitudinal muscle include:
    - The muscle appears in the upper (superior) portion of the tongue.
    - It extends longitudinally along the length of the tongue, from the base to the apex.
    - On T2w MRI, the muscle is hypointense and appears darker than surrounding tissues.
    - It starts from the most anterior portion of the palate, which can be denoted on T2w images as a hyperintense band above the tongue.
- Start in the sagittal view and locate a good slice to begin – look for a slice with a clear view of the palate, the tongue, and the space (void) between the tongue.
- In most subjects, there is a void of air between the tongue and palate in most slices (the space where void of air is the oral cavity proper).
- The superior longitudinal muscle continues to the transverse/vertical muscle below it.

### a. Sagittal View

1. Begin with the sagittal view for segmentation.
2. Start more towards the central sagittal slice. This slice should provide a clear view of the muscle to be segmented. It is easier to start here and then label laterally, rather than start at the lateral ends. Look for a slice that shows the palate and the tongue, separated by a space, the oral cavity proper (labelled purple).
3. Identify the superior longitudinal muscle - on a T2-weighted image, it appears as a hypointense band running across the top quarter of the tongue. The muscle begins at the anterior-most portion of the palate, which you can spot on T2w as a hyperintense band above the tongue (labelled dark blue).
    a. Above the superior longitudinal muscle, there may be a void of air (oral cavity proper) (labelled purple), between the tongue and the palate in some slices, which should not be labelled the superior longitudinal muscle. This part is more hypointense usually.
4. Below the superior longitudinal muscle is the transverse/vertical muscles (labelled green). The transition is where the hypointense region either becomes more speckled/hyperintense, or there is a hyperintense line separating them.

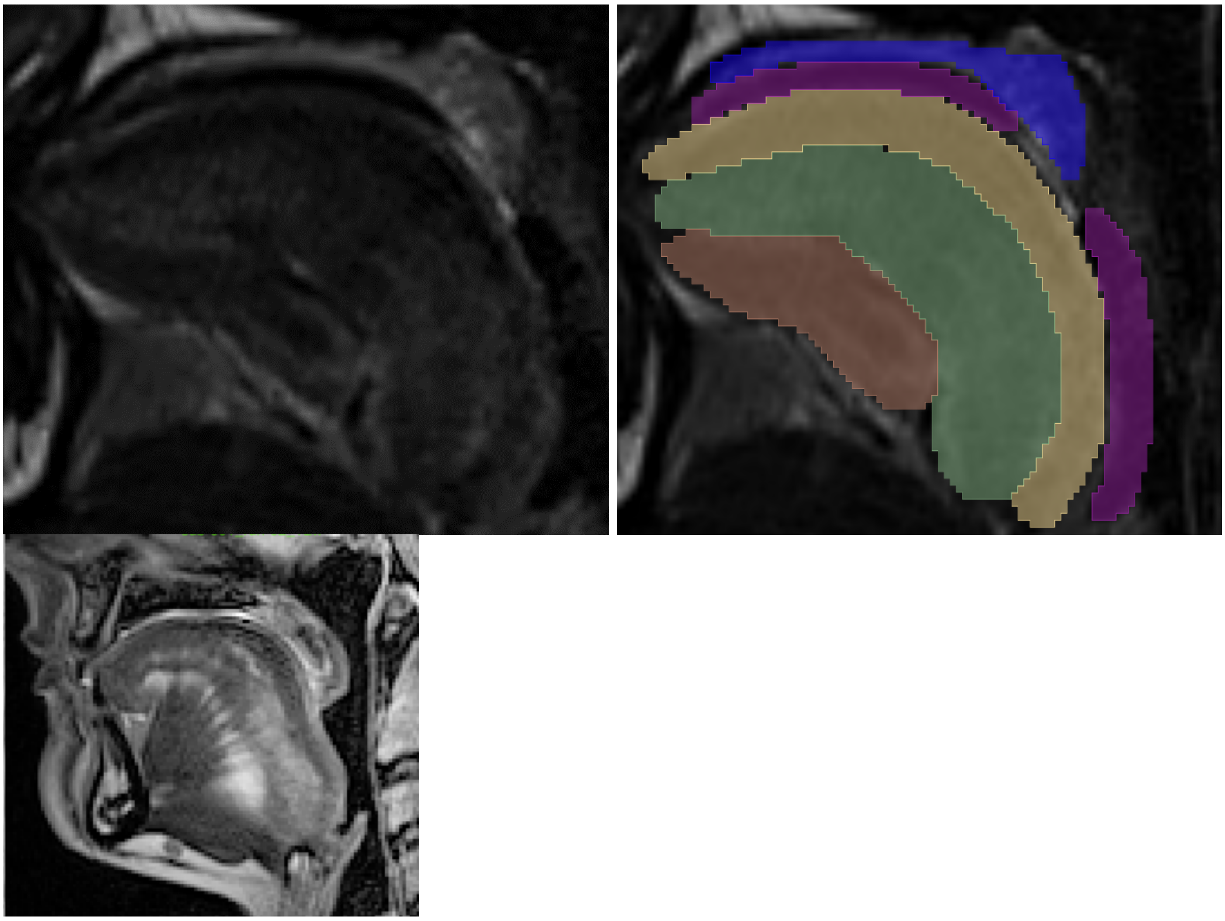

Figure 1: Sagittal view of the tongue and labels (T2w). Look for the hypointense band (bottom) – windowing of the image is important.

### b. Axial View

After the sagittal view, proceed to axial view.

1. The superior longitudinal muscle (yellow) will be more posterior (towards the back) compared to the transverse/vertical muscle (green).
    a. Notice on the sagittal view how the superior longitudinal muscle arcs downwards behind transverse/vertical muscle.
    b. It is also typically more of a “W” shape here.
    c. As you move towards the superior ends (scrolling upwards), the muscle will become a larger sheet (given the top of the tongue is just the superior longitudinal muscle).
    d. This is shown in the second set of images. (Fig 2)

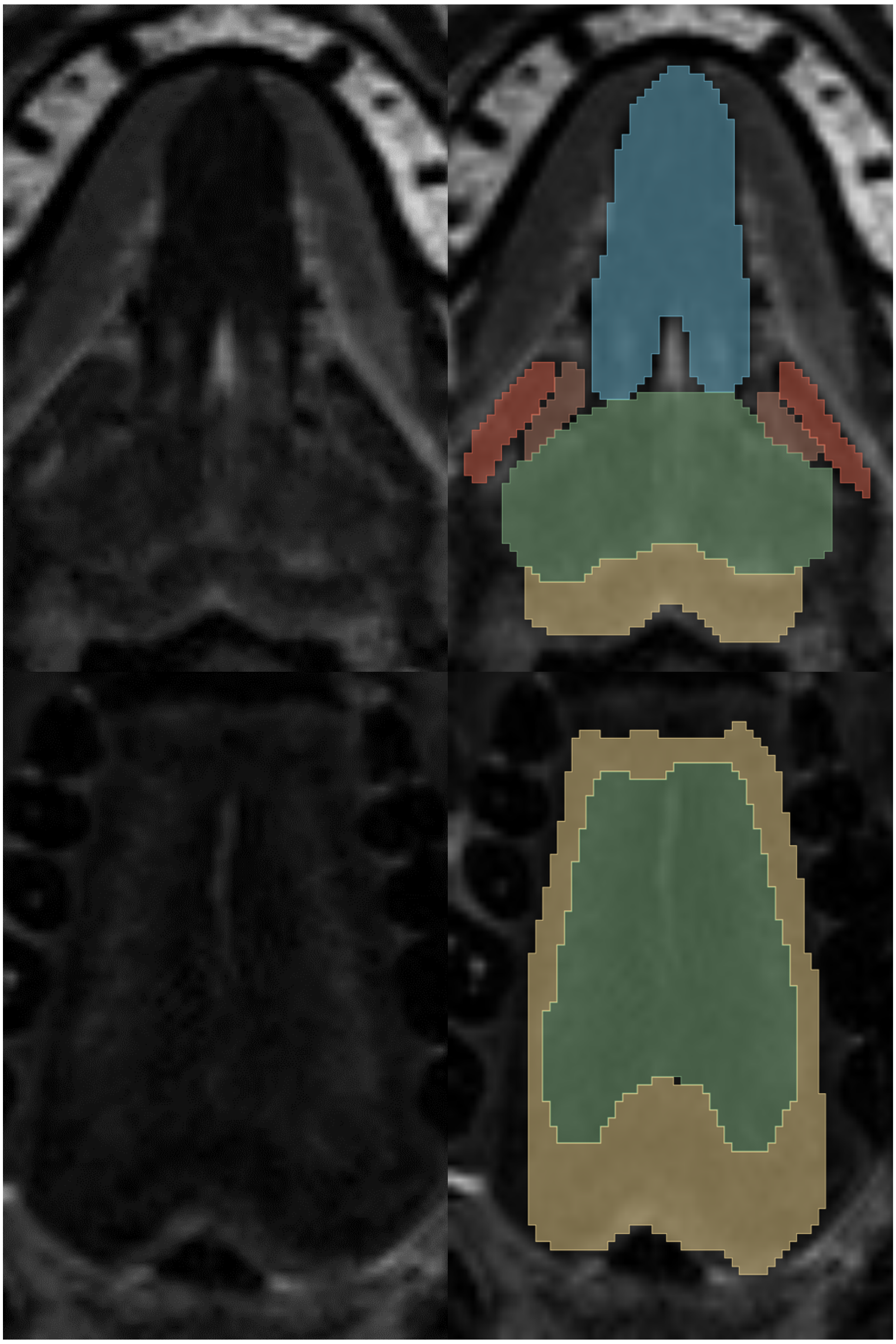

Figure 2. Axial view of a T2w image of the tongue, showing the distribution of the superior longitudinal and beginning of the transverse/vertical muscle.  Note the W shape

### Coronal View

After the axial view, proceed to coronal view.

1. The superior longitudinal muscle (yellow) will be more superior compared to the transverse/vertical muscle (green).
    a. It is also typically more of a “M” shape here.
2. Notice again the air in the oral cavity proper above the tongue (purple), and the palate (dark blue).

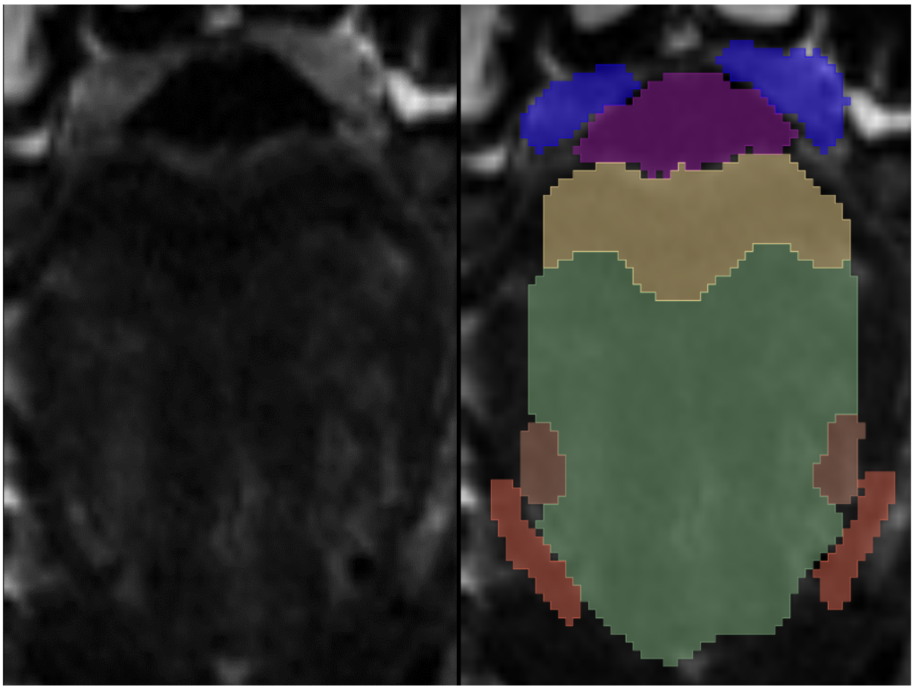

Figure 3. Coronal view of the tongue musculature on T2w image. Note the M shape of the superior longitudinal and transverse/ vertical.

### 2) Transverse/vertical muscle (labelled green)

#### General overview

- The overall location of this muscle is below the superior longitudinal muscle.
- There is an obvious curvature gradient that can be followed for both the superior longitudinal and transverse/vertical muscles.
- On the sagittal view, search for a speckled and hyperintense band below the superior longitudinal muscle. The transverse/vertical muscle has a similar form to the superior longitudinal muscle, where it curves as it moves more posteriorly.
- When there is difficult delineation, the best technique is to revert to the previously labelled slice and copy the curvature from there as a reference.

#### a. Sagittal View

1. Starting with the sagittal view, find the transverse/horizontal muscle (green) by locating the speckled, more hyperintense region below the superior longitudinal muscle (yellow).
   a. There is an obvious curvature gradient that can be followed for both the superior longitudinal and transverse/vertical muscles.
   b. Sometimes in more difficult scans the muscle can be less speckled or clearly delineated, and the only identifiable transition points between muscles is a thin hyperintense line (labelled in grey; between transverse/horizontal muscle <-> superior longitudinal muscle and transverse/horizontal muscle <-> genioglossus).

b. The inferior border is shared with either genioglossus (light blue) medially (as shown in first set of images) or inferior longitudinal muscle (salmon) laterally (as shown in second set of images).

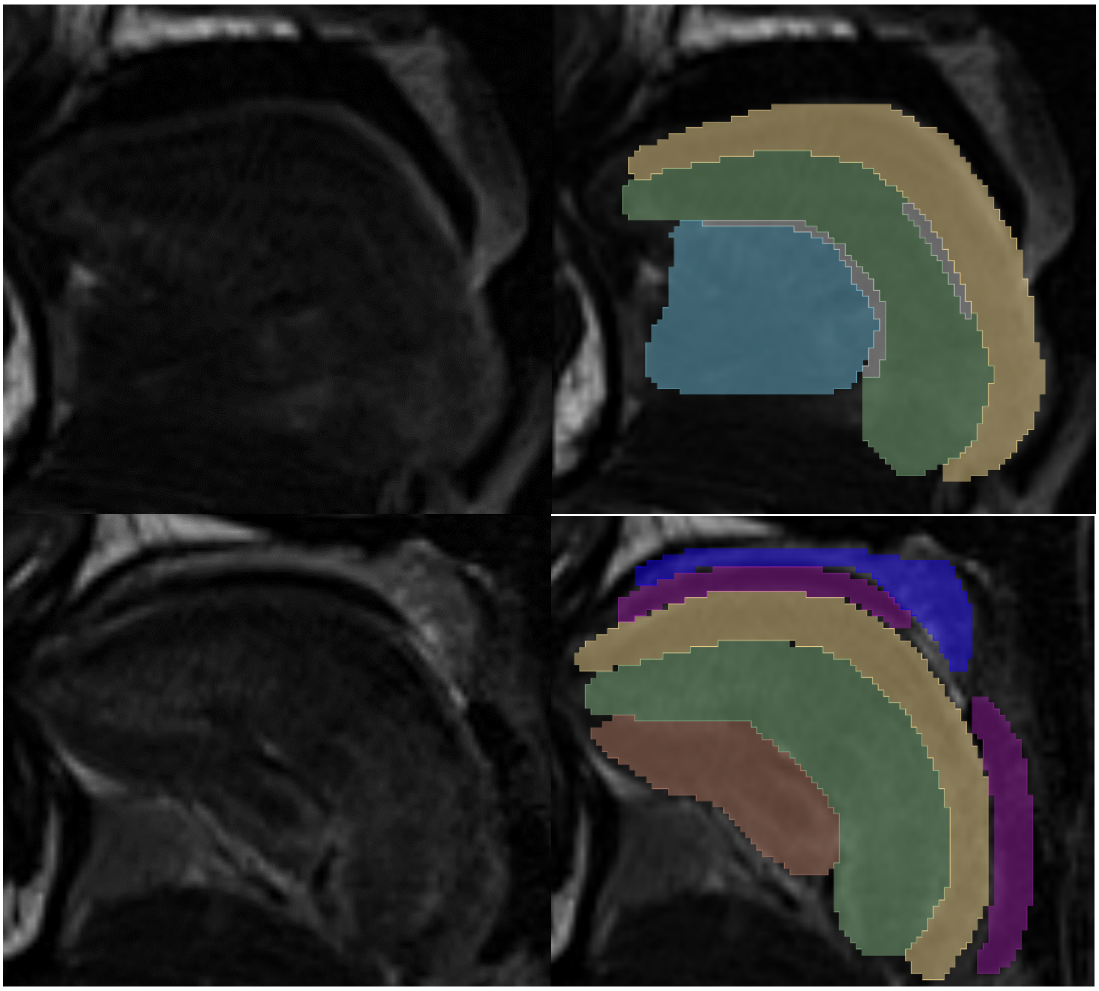

Figure 4. Sagittal view of the tongue and labelled muscles, showing the gradient and angle of the palate, and how these are mirrored in the superior longitudinal and transverse/vertical muscles.

### b. Axial View

1. In the axial view, the transverse/vertical muscle (green) is best observed in the middle third of the tongue, extending laterally from the midline towards the edges of the tongue.
    a. The muscle is more anterior to the superior longitudinal muscle (yellow). The inferior longitudinal muscle (salmon) and genioglossus (blue) are both more anterior to transverse/vertical muscle.

b. Note the bright thin hyperintense (brighter) band that outlines the transverse/vertical muscle.

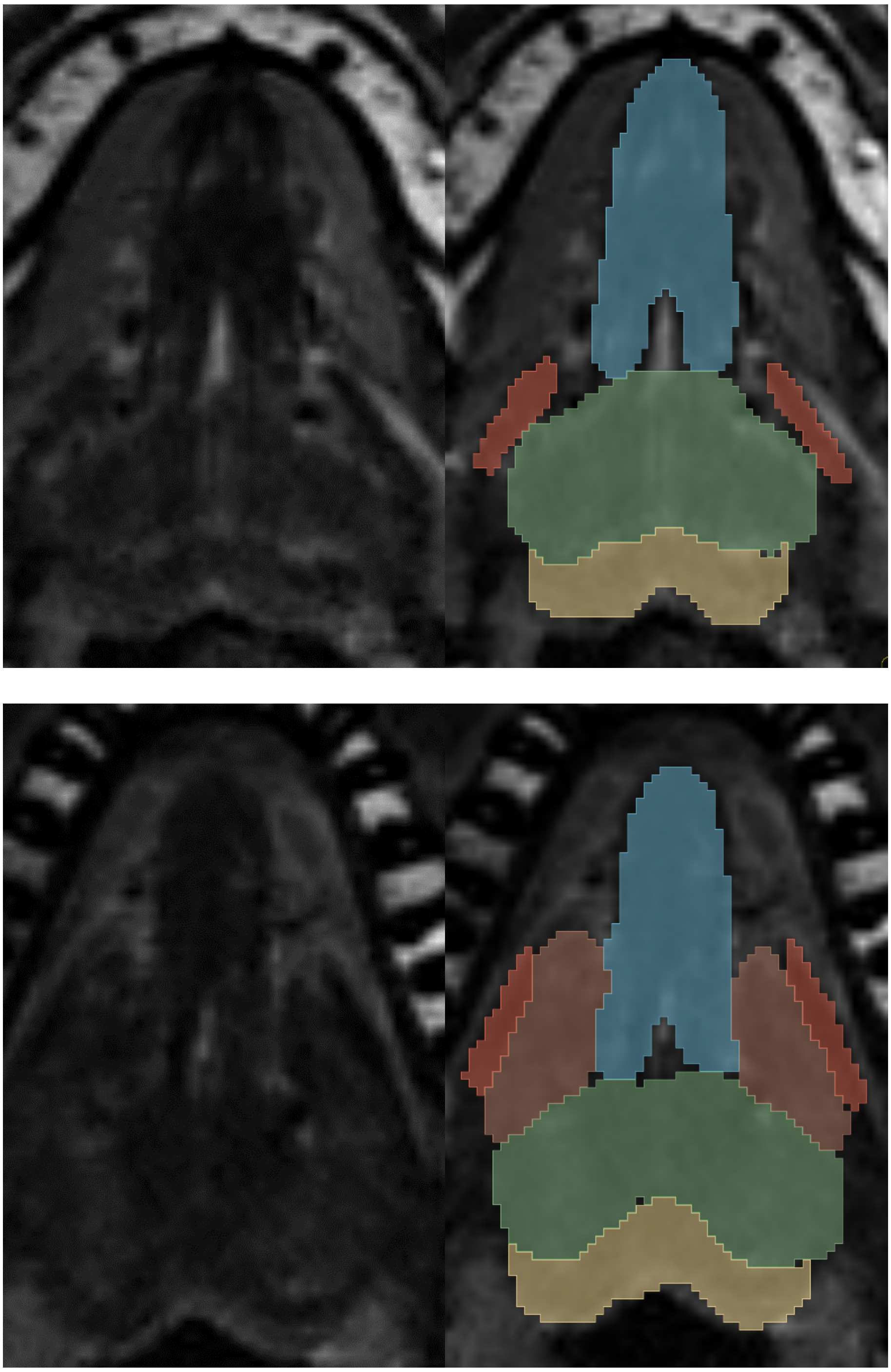

Figure 5. Axial view of the tongue on T2w slices. Note the hyperintense lines that separate the transverse/vertical muscles from the surrounding muscles.

### c. Coronal View

In the coronal view, the same principles apply.

- The transverse/vertical muscle (green) is below the superior longitudinal muscle. Towards the posterior tongue (Fig 6, top), it takes up a larger portion of the tongue, while anteriorly (Fig. 6, bottom), there is less mass.

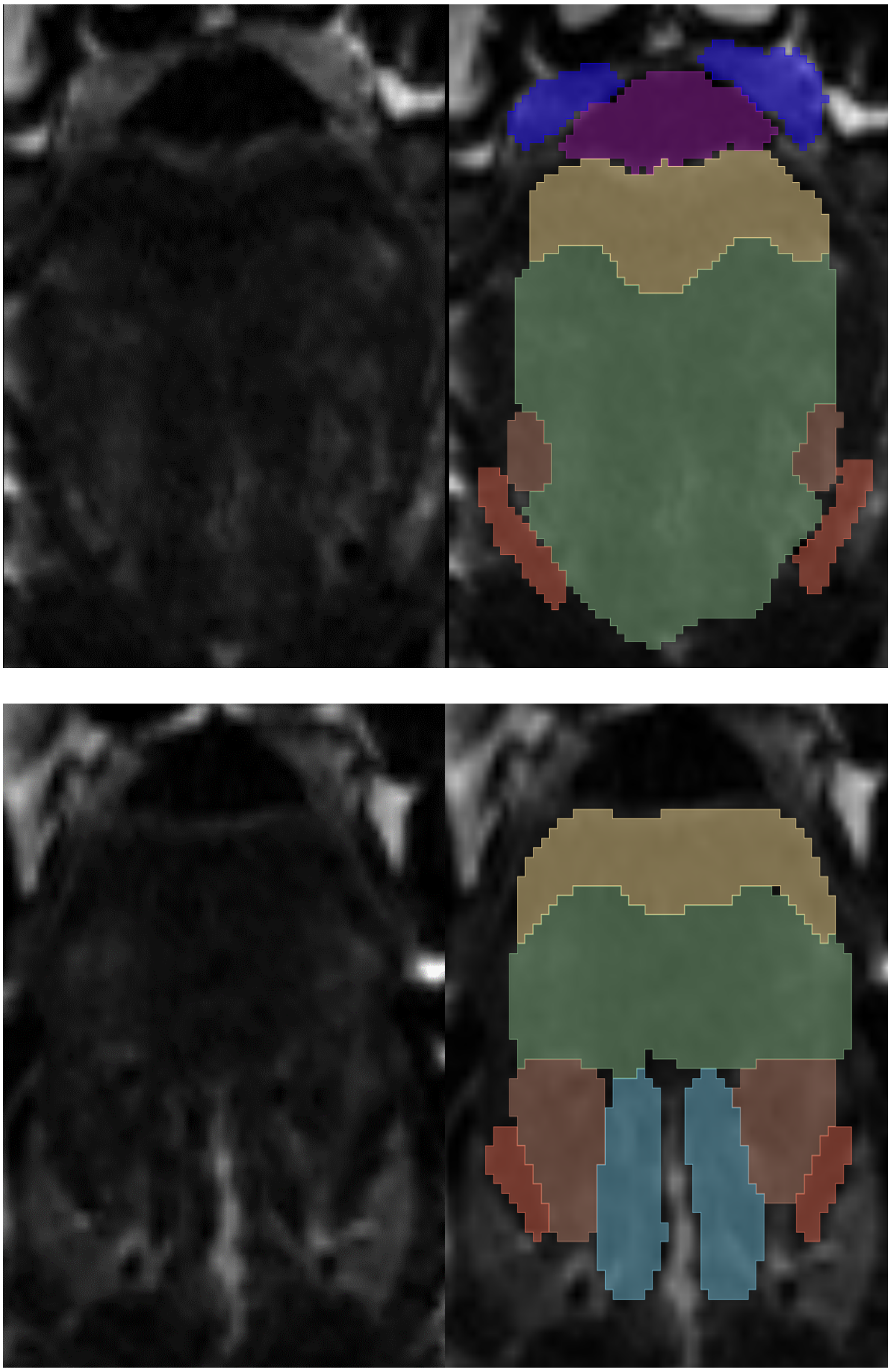

Figure 6: the coronal view of the tongue shows the same M shape as the longitudinal muscle and follows the tongue about halfway down. It is important at this point to adjust the windowing of your viewer to ensure the contrast between muscles is visible.

### 3. Inferior Longitudinal Muscle (labelled salmon)

#### General overview:

- There are three main muscles in the inferior region of the tongue below the transverse/vertical muscle. These are the genioglossus, the inferior longitudinal, and the hyoglossus muscles.
- The inferior longitudinal muscle is inferior to the transverse/vertical muscle towards the lateral edges of the tongue (between genioglossus and hyoglossus).
- It is typically more hypointense than transverse/vertical muscle.

#### Sagittal View

1. Begin with the middle sagittal slice.
2. From this point, transition halfway towards the lateral extremity of the tongue.
    a. You should be able to identify the inferior border of the transverse/vertical muscle (labelled green), this is the transition point to the superior portion of the inferior longitudinal muscle (labelled salmon).
    b. The inferior longitudinal muscle mirrors the curvature of the superior longitudinal muscle (labelled yellow) and the transverse/vertical muscle (labelled green).
    c. The muscle typically appears hypointense on T2w images.
3. Once identified, follow the muscle around towards the sublingual (salivary) gland (labelled pink in Figure 7), located directly inferior to the muscle.
    a. The salivary gland provides a clear landmark as it exhibits a brighter intensity gradient than the surrounding tissues.
    b. Note that as you move more lateral, hyoglossus (labelled red) will appear in the images and insert into the inferior longitudinal muscle medially. This is shown in the second set of images.

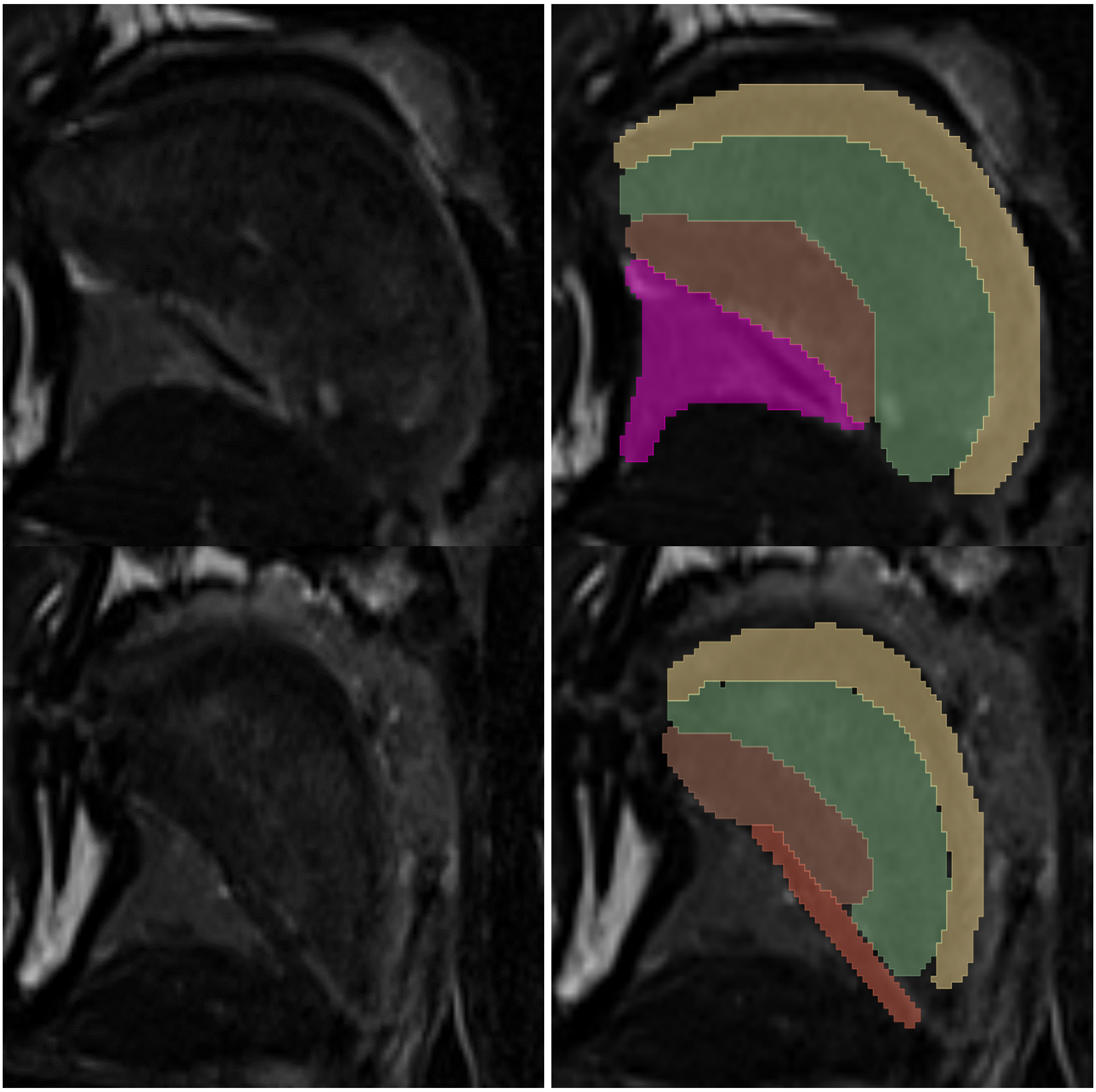

Figure 7: Sagittal view. Note the salivary gland (pink) and the salmon inferior longitudinal.

### b. Axial View

After initial identification and segmentation on the sagittal view, move to the axial view.

- The inferior longitudinal muscles are more anterior (in front of) the transverse/horizontal muscle in most slices.

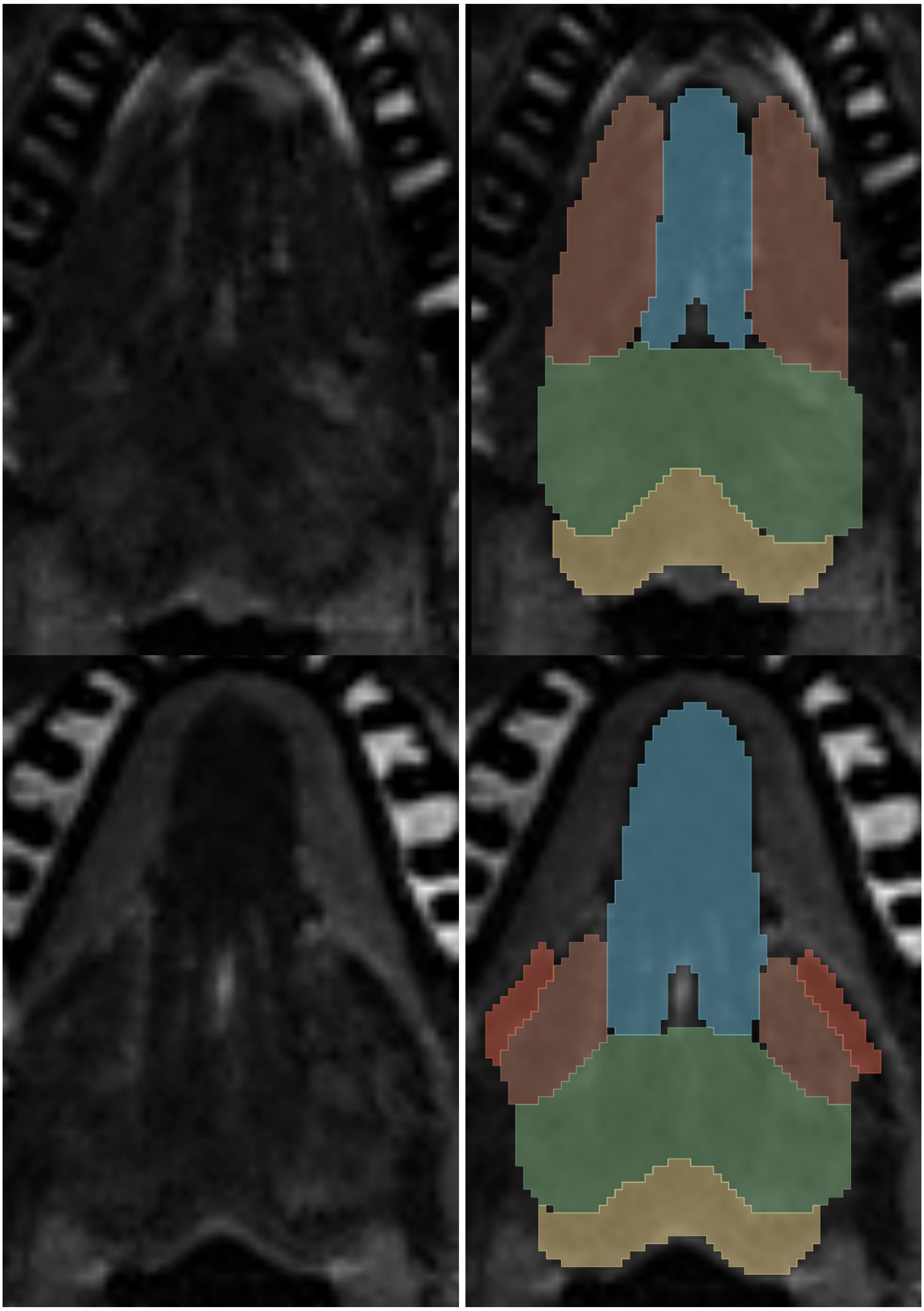

Figure 8: axial view highlighting the inferior longitudinal (salmon), which is bordered by the hyoglossus (red) in the more inferior slices.

Another helpful landmark is genioglossus (labelled blue). It will be helpful to revise the label of inferior longitudinal after labelling genioglossus as well. The inferior longitudinal muscles surround genioglossus, typically having a hyperintense line that separates the two muscles. In the superior portions of the tongue, the height on the image seen in the first set of images is similar between genioglossus and inferior longitudinal muscle. However, as you transition

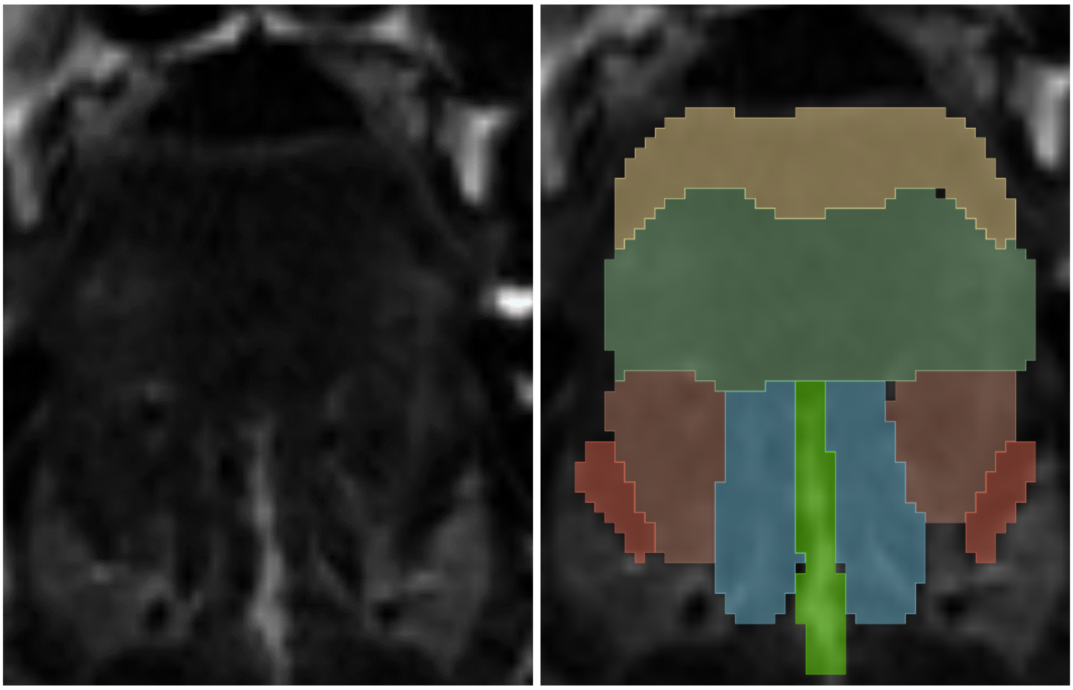

Figure 9: coronal view showing the location of the inferior longitudinal. Note the shape and borders – the hyoglossus appears about 1/3 of the height and below. Note that the bright band in the middle is the tongue septum (bright green).

more inferiorly on the axial view, the inferior longitudinal muscle becomes less visible, replaced by a thin, darker band of muscle denoting the hyoglossus (labelled red). This is highlighted by the comparison between the first and second set of images.

#### c. Coronal View

In the coronal view, three main muscles become apparent in the inferior region of the tongue, namely (from medial to lateral) the genioglossus (blue), the inferior longitudinal (salmon), and the hyoglossus (red) muscles. These muscles appear and disappear at various stages as you transition and scroll from the middle slice to the more anterior and posterior slices. Note that the bright band in the middle is the tongue septum (bright green).

### Genioglossus (labelled blue)

#### General overview

- Best labelled in the axial view, where you can look for the inverted hypointense V shape at the anterior tongue.
- Inferiorly, there is geniohyoid, and superiorly, there is the transverse/vertical muscle.
- Moving from medial to lateral is tongue septum -> genioglossus -> inferior longitudinal -> hyoglossus.

#### a. Axial View

For the genioglossus, start with the axial view since this muscle is best visualized in this plane.

On both T1w and T2w images, you're looking for an inverted V-shaped structure (blue) that starts posterior to the mandible and towards the chin. This structure is the genioglossus muscle. On T2w images, the genioglossus muscle will appear darker, or hypointense. The reason it is an inverted V-shaped structure is because the tongue septum is not labelled (the hyperintense structure between the two ends of genioglossus posteriorly on the axial view).
As you progress towards the inferior slices, you will notice a semicircular shape form on the lateral sides of the muscle. This denotes the transition to the geniohyoid muscle inferiorly (labelled purple in the second set of images).
Moving superiorly, the genioglossus muscle starts blending into the transverse/vertical muscle, which you have already labelled in the sagittal view. The muscle boundary may become less defined as these muscles are closely packed and overlap. Note that superiorly, genioglossus is surrounded laterally by the inferior longitudinal muscles (shown in the third set of images).

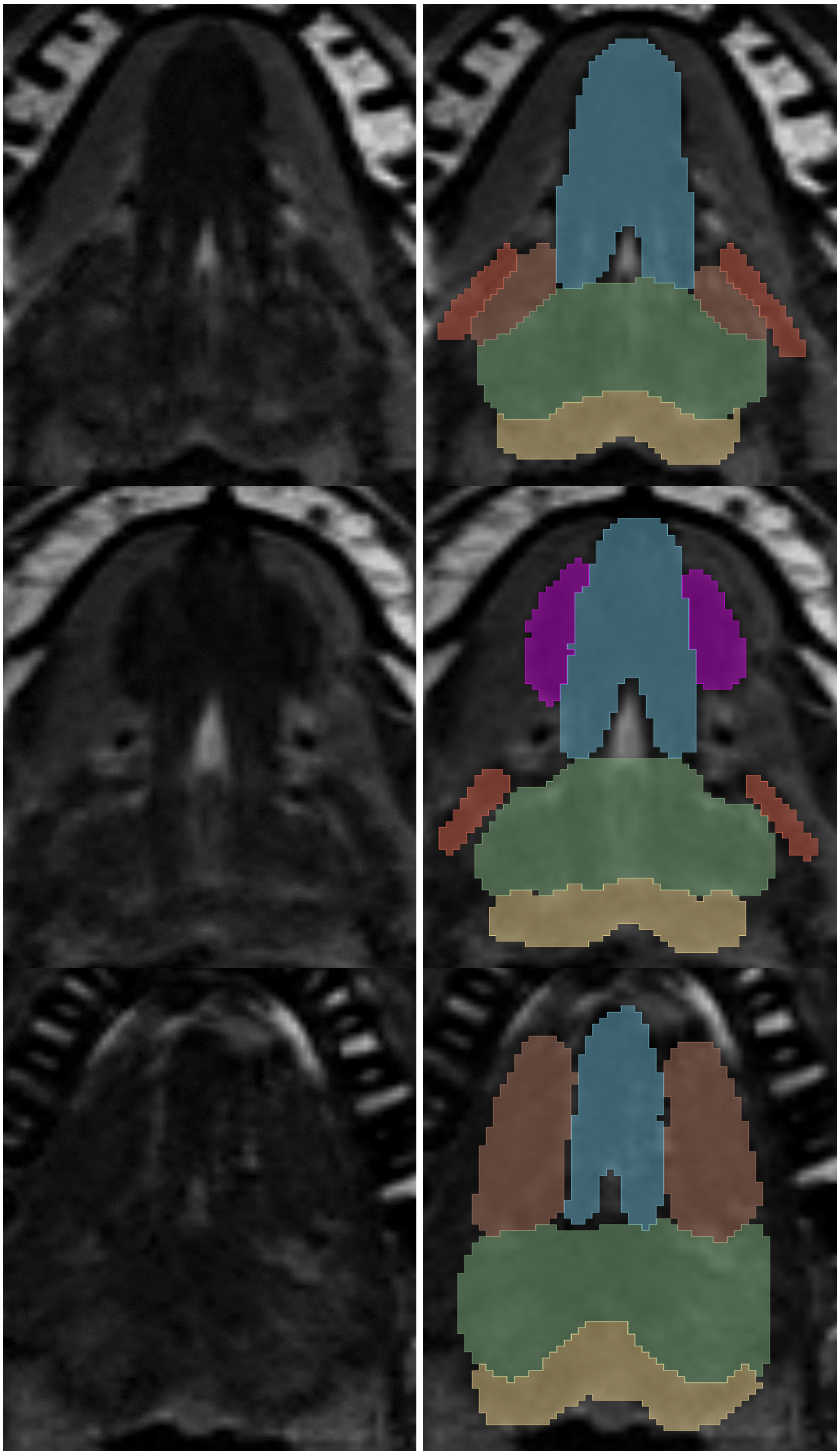

Figure 10: axial views of the genioglossus. Note thev shape, avoiding the septum.

### b. Sagittal View

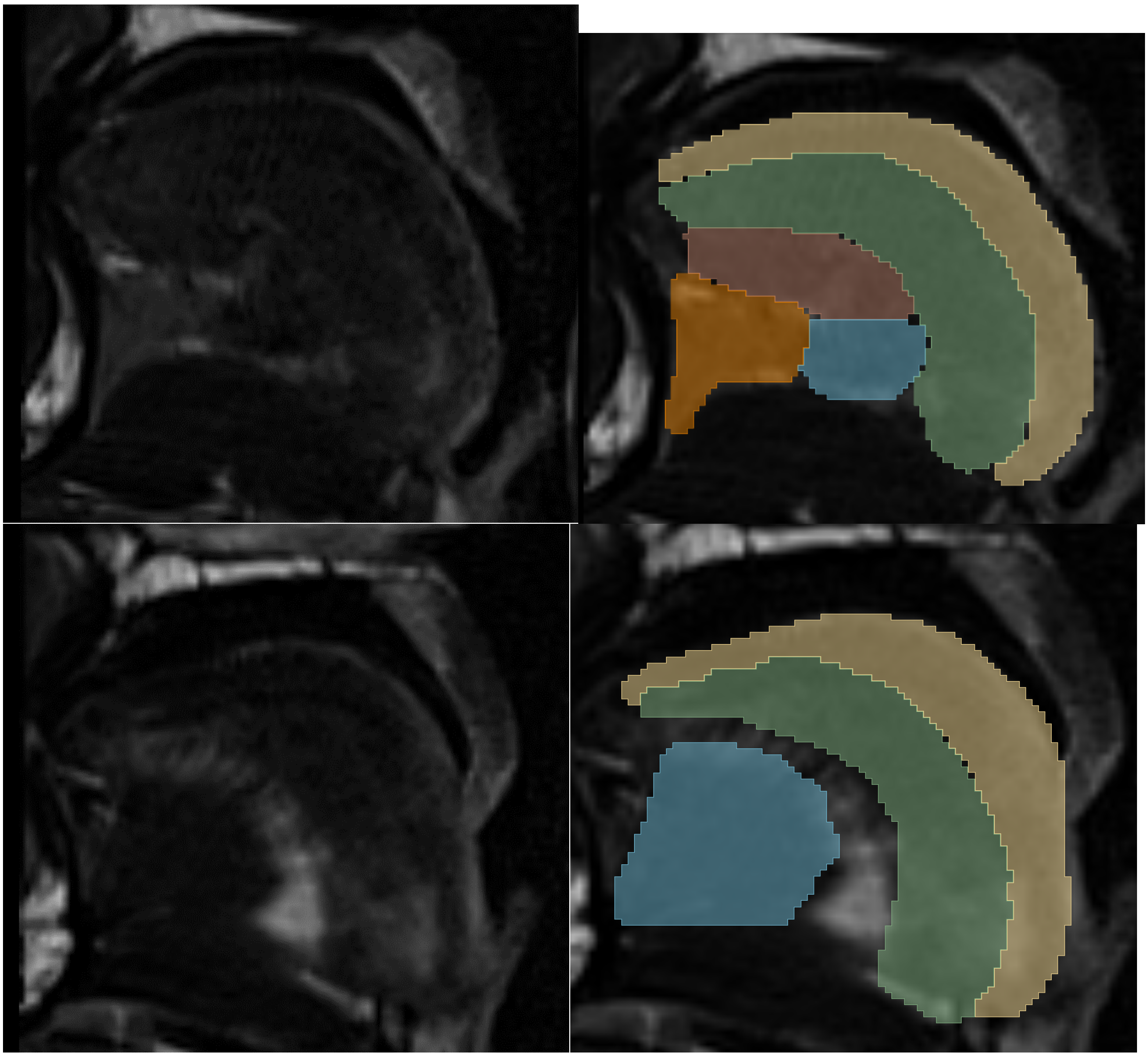

Figure 11: sagittal views of the genioglossus. In this plane, the key landmarks to note are the sublingual gland (labelled orange) and the tongue septum (not labelled in the key images). The sublingual gland is anterior to genioglossus (seen in the more lateral slices, first set of images) and the tongue septum is posterior to genioglossus (seen in the more medial slices, second set of image; wedged between genioglossus in blue and transverse/vertical in green). Note that both key structures are more hyperintense than the muscles in T2w images.

### c. Coronal View

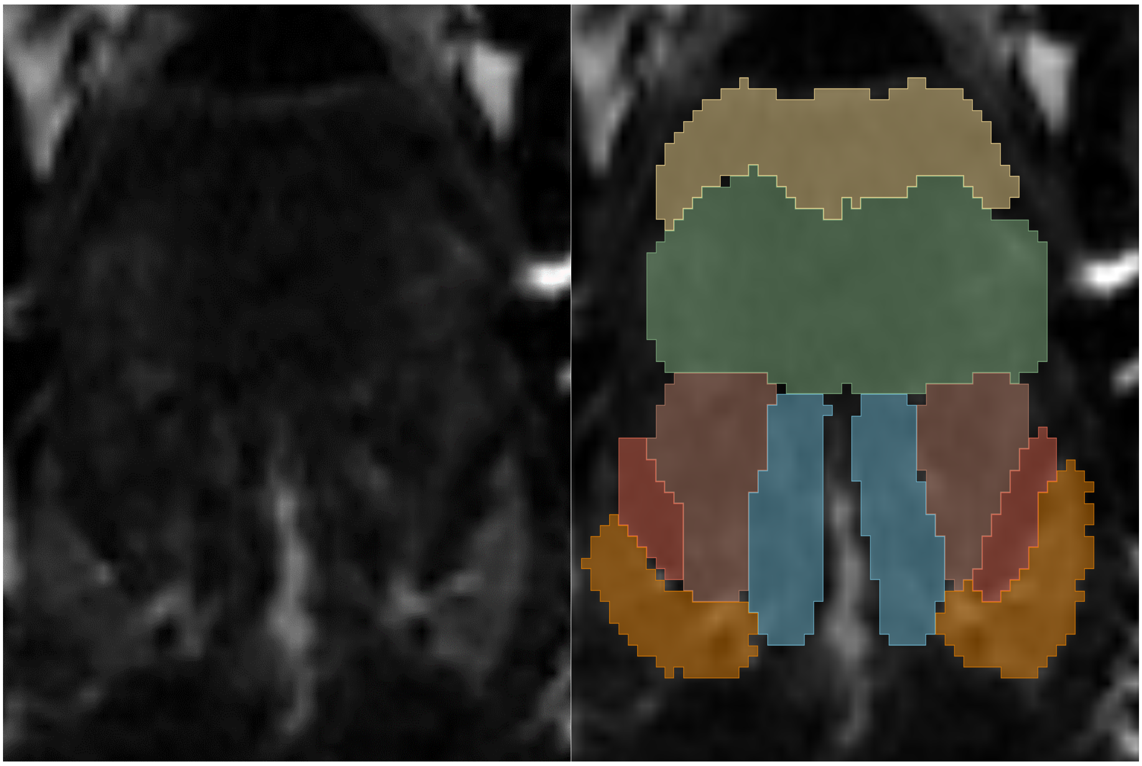

Figure 12: Coronal view.
The same principles apply in this view. Transverse/vertical muscle superiorly, and inferior longitudinal muscle laterally. Note again the hyperintense line between genioglossus, the tongue septum. From medial to lateral, the structures are as follows. Tongue septum (not labelled), genioglossus (blue), inferior longitudinal muscle (salmon), hyoglossus (red), sublingual salivary gland (orange). As you move anteriorly, the other muscles surrounding genioglossus become smaller (hyoglossus disappears, then inferior longitudinal muscle disappears; as you move closer to the tip of the tongue).

### Hyoglossus (labelled red)

#### General instructions

- The best view to label this muscle is axial, where the hyoglossus is a uniform, roughly rod-shaped structure with rounded edges.
- The most consistent way to find hyoglossus is by identifying a characteristic group of surrounding structures/landmarks. These are the sublingual gland, submandibular gland, submandibular duct, mylohyoid and the transverse/horizontal muscle.
- The hyoglossus transitions/inserts into the inferior longitudinal muscle superiorly.
- Bottom (inferior) can be difficult to delineate – the shape of the muscle does not expand in the more inferior slices, where it connects to the styloglossus muscle, and rather stays in its characteristic rod shape.

a. Axial View

The hyoglossus can be identified as a dark, or hypointense band. Its characteristic morphology in the axial plane is roughly rod-shaped with rounded edges, giving it a distinctive appearance.

On the axial view, begin your search by finding the characteristic group of surrounding structures/landmarks. These are the sublingual gland (orange), submandibular gland (bright green), submandibular duct (dark blue), mylohyoid (yellow) and the transverse/horizontal muscles (dark green). You should see two hypointense oval/rod shaped structures (the two muscles of hyoglossus and mylohyoid) with a bright hyperintense line between them (which is the submandibular duct). Note that hyoglossus is the medial black rod, while the mylohyoid is the lateral one. Above and below the hyperintense line should be two grey structures (with an intensity that is between that of the duct and the muscles). These are the sublingual and submandibular glands above and below, respectively. Medially, the hyoglossus abuts the transverse/horizontal muscle.

Start by labelling the hyoglossus in the axial slices where it is clearly visible and maintain consistent anatomical boundaries as you navigate through the axial plane. Continue following the hyoglossus superiorly through the axial slices until it merges inferior longitudinal muscle. Inferiorly, it is very difficult to ascertain the exact boundary. Typically, it will reach a similar end point as when the rest of the tongue ends (i.e., When transverse/vertical muscle finishes as well).

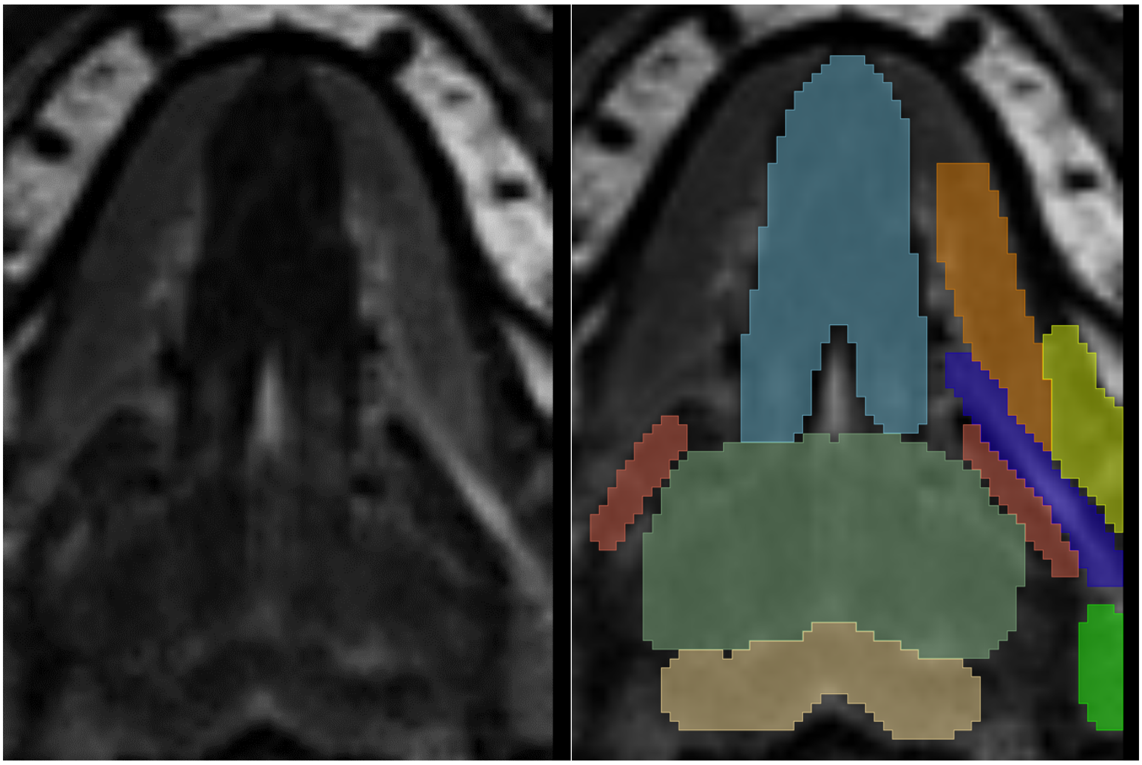

Figure 13. Axial view of the tongue – note the hyoglossus (red) in the left image, clearly visible at the lateral edges of the tongue.  This landmark is also useful for finding the border of the Transverse/vertical muscle.

### b. Sagittal and Coronal Views

The sagittal and coronal views provide complementary perspectives for hyoglossus muscle segmentation. Although the muscle is best visualized and labelled in the axial view, sagittal and coronal planes can help refine the segmentation, especially at the borders where the muscle transitions to neighbouring structures.

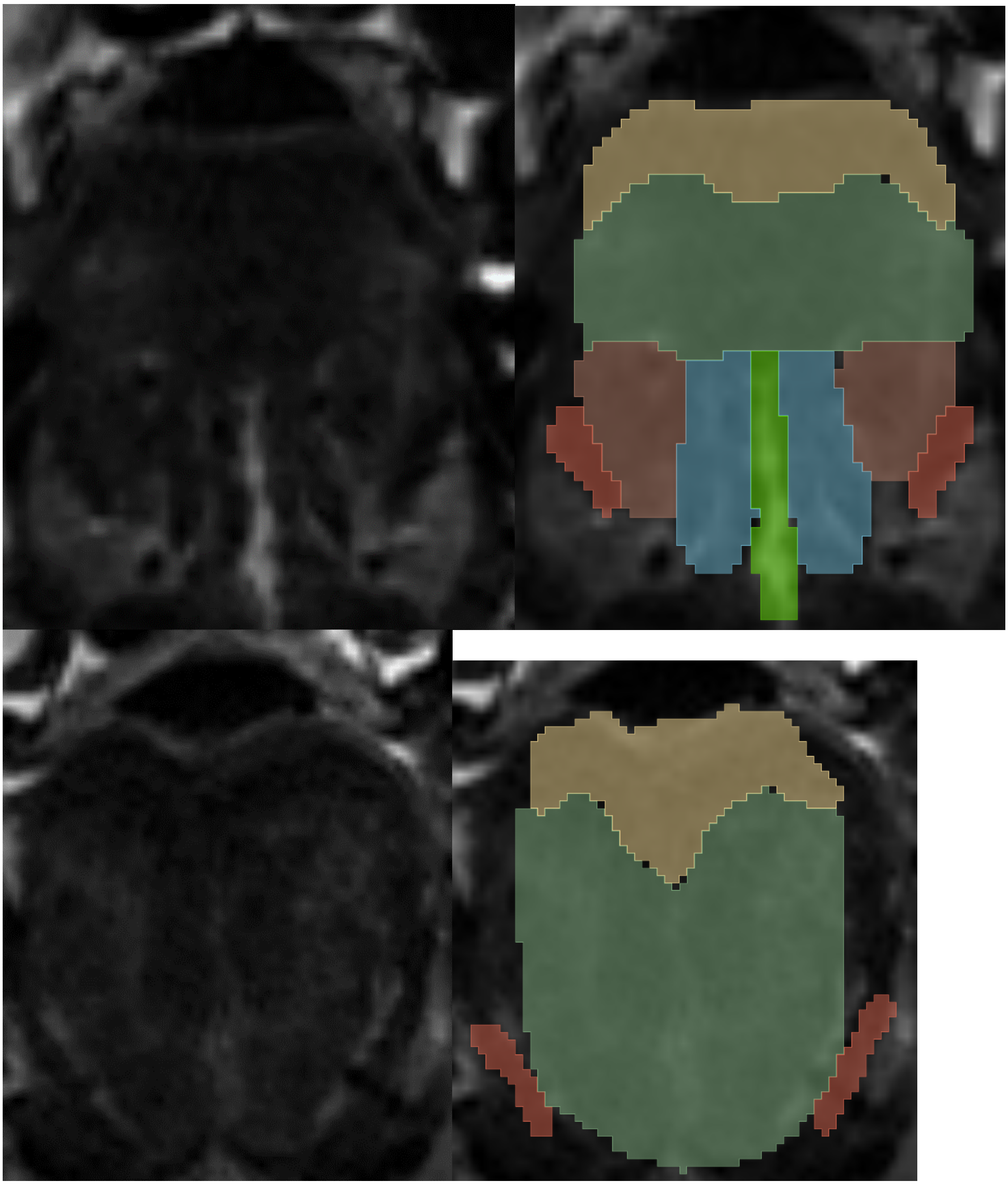

Figure 14: In coronal view, the hyoglossus muscle is found as the most lateral structure. In the second set of images below, the order of the muscle from medial to lateral is as follows. Tongue septum (note that this is labelled bright green in this instance), genioglossus (labelled in blue), inferior longitudinal muscle (labelled in salmon), and lastly hyoglossus (labelled in red). The third set of images shows you the muscles as you move more posteriorly. Note how the inferior boundary of hyoglossus is very difficult to ascertain, however in general should end similarly to the transverse/horizontal muscle. The superior boundary is again difficult to determine. The best method is to use guidance from previous slices. Superiorly, hyoglossus intertwines with styloglossus.

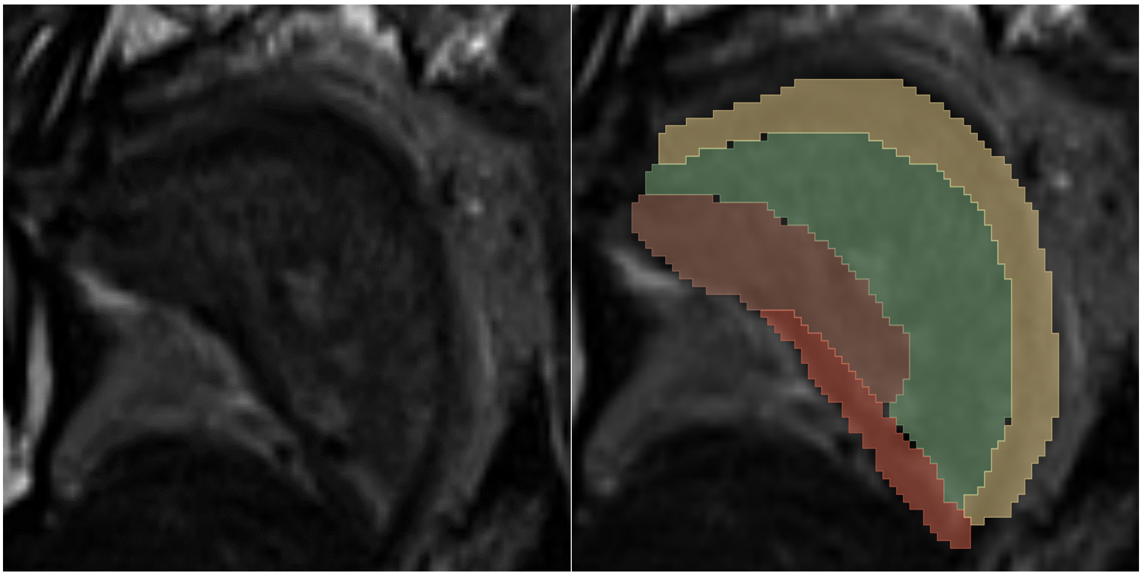

Figure 15: In the sagittal view, the hyoglossus muscle appears as a thin, elongated structure located laterally. It is inferior to the other tongue muscles, inserts into the inferior longitudinal muscle, and typically runs the length of the other tongue muscles
