## Appendix Two for "Segmentation of the Human Tongue Musculature Using MRI: Field Guide and Validation in Motor Neuron Disease"

### Appendix Two for Segmentation of the Human Tongue Musculature Using Magnetic Resonance Imaging: Field Guide, Atlas, Tool, and Datasets.

#### Section One: Intra-oral cavity measurements.

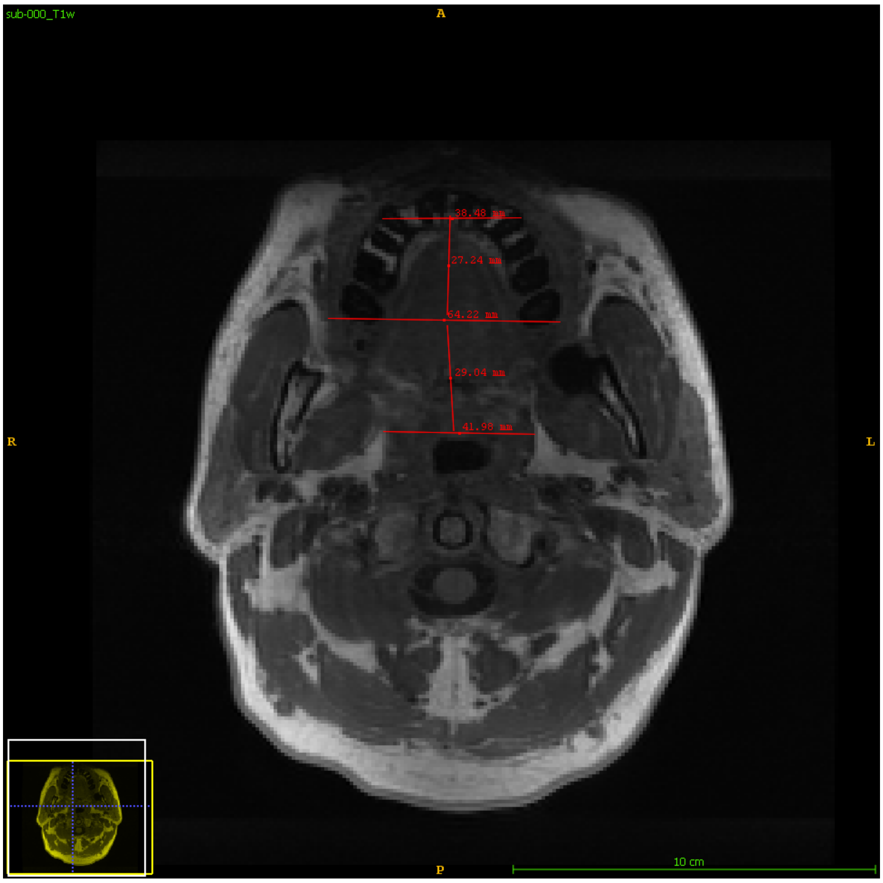


Appendix T2o Figure One: The measurements of the frustums in an axial view of the IOC

#### Frustum/ Kegelstumpf(s) of the oral cavity

##### 1. Overview

We measure two truncated cones (or kegelstumpf) to get the overall Intra Oral Cavity (IOC) volume.

Our assumption for measuring the IOC is:

1. The ratio of diameters between the long axis of the top and bottom (midline) is equal to the ratio of the short axis top and bottom short axis measurements.

#### 2. Identify the Axial Slice for Radius (r) Measurement:

1. Go to the middle sagittal slice, where you can view the brain stem or the septum of the tongue. Draw the midline (the largest line from the palate to the base of the hyoid)
2. Switch to an axial view, note the midline. Draw an intersecting line from the left to right of the mouth, from the midline (at its widest point). You now have the midline mapped in the axial and sagittal planes.
3. Move superiorly and identify the top jaw and place your cursor on the teeth. Looking at the axial view, note where your cursor is placed.
4. Take the measuring tool, and measure between the left and right upper incisors (from the outer edge of the incisors to the other side. This is the top of the first truncated cone.
5. Connect the incisor line to the midline in the axial plane. This is the height of the first cone
6. Identify the parapharyngeal fat near the oesophagus. You may need to move superior or inferiorly to your current position, noting the midline in the axial plane.
7. Connect the two sides of the parapharangeal fat. This is the top of the second truncated cone.
8. Again, connect the top of the cone and the midline on the axial slice. This is your second height. Ensure the measurements are accurate on each slice view. Move a few slices around on each view to ensure consistency. We noticed that the measurements may vary greatly depending on the slice, so it is best to take a few measurements and average.

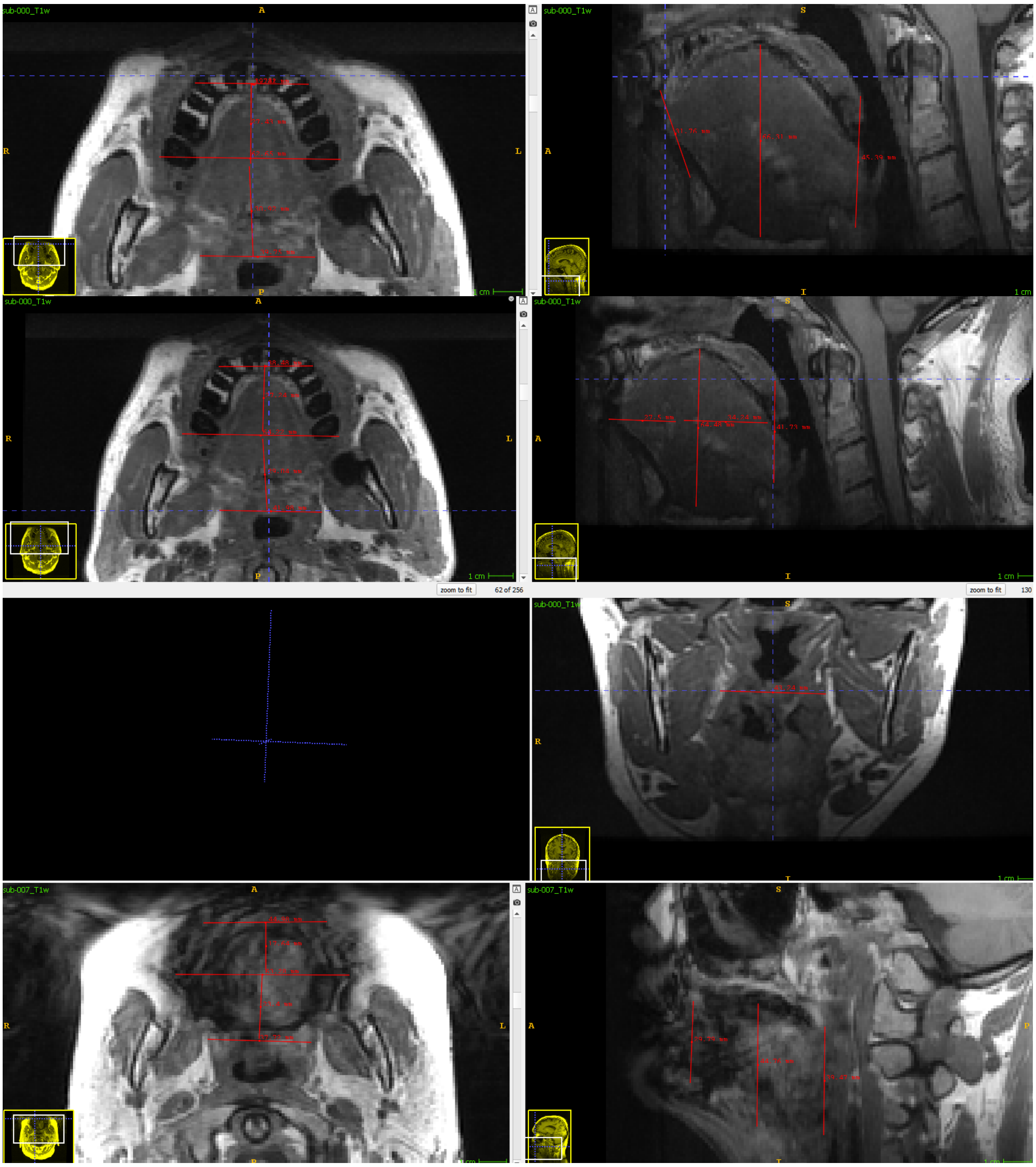


Appendix Two Figure Two. (Top two rows) Good quality images of the tongue and the different measurements that are required to calculate the IOC.
Bottom 2 rows: Poor quality tongue images require an estimation of the boundaries and should likely be excluded from analysis.

### Calculate the Volumes:

#### 12. Calculate Oral Cavity Volume:

Need to make two kegelstumpfs with elipses instead of circles. (truncated elliptical cone – frustum).
The volume of the frustum within the right elliptical cone can be expressed as:

$$V = \frac{\pi}{3} \times (H \times r_{SA-b} \times r_{LA-b} - j \times r_{SA-t} \times r_{LA-t})$$ *Eq. 1*

Eq. 1:

Where *j*: The height of the topmost cone (that is removed) before truncation, H: The height of the entire elliptical cone (before truncation), $r_{SA-b}$: The shortest radius of the base of the frustum, $r_{LA-b}$: the longest radius of the bottom base of the frustum, $r_{SA-t}$: the shortest radius of the top of the frustum, $r_{LA-t}$: the longest radius of the top of the frustum and assuming $r_{SA-b}/r_{LA-b} = r_{SA-t}/r_{LA-t}$.

As *H* and *j* are unknown, due to the biology not existing past the top of the frustum, we derive *H* as a function of the radii and the measured height h of the fustrum, which is the portion of the cone remaining after truncation, as follows

1) Solve for *j* = the height of the topmost cone

$$j = H - h$$ *Eq. 2*

2) Based on intercept theorem for the short axis a the top of the frustrum $r_{SA-t}$

$$r_{LA-t} = \frac{r_{LA-b} \cdot j}{H}$$ *Eq. 3*

3) Therefore, we can solve for H:

$$r_{LA-t} = \frac{r_{LA-b} \cdot (H - h)}{H}$$

$$r_{LA-t} = r_{LA-b} \cdot \left(1 - \frac{h}{H}\right)$$

$$r_{LA-t} = r_{LA-b} - \frac{r_{LA-b} \cdot h}{H}$$

$$r_{LA-b} - r_{LA-t} = \frac{r_{LA-b} \cdot h}{H}$$

$$H \cdot (r_{LA-b} - r_{LA-t}) = r_{LA-b} \cdot h$$

4) And therefore *H:*

$$H = \frac{r_{LA-b} \cdot h}{(r_{LA-b} - r_{LA-t})} \qquad \textit{Eq. 4}$$

We measure the two radii of the base (larger ellipse) of the frustrum and propagate the ratio of the smaller radius (I.e., *b*) to the top frustrum (the smaller ellipse, i.e., *d*). This minimises the error propagation.

#### Section Two: Quality control

**Appendix Two, Table One.**

Segmentation and MRI rating criteria for tongue MRI segmentation. Each of these criteria were adapted from [1] and [2].

| **Quality Rating** | **Segmentation Description** | **MRI Description** |
|---|---|---|
| **1** | Unusable Quality: Either completely unusable segmentations or labels that do not fit the anatomy. Fail | Unacceptable Quality: This includes scenarios where the tongue is not visible, there are large artifacts obscuring the tongue, extreme motion artifacts are present, or there's a complete lack of contrast or signal. |
| **2** | Poor Quality: Missing large sections of the required labels or having labels in incorrect locations. Alternatively, this level can also include many (5+) small errors in segmentation. Fail | Poor Quality: Characterized by smaller artifacts like those described in 1, or minor motion artifacts. However, the tongue is still identifiable despite these issues. (fail or check) |
| **3** | Moderate Quality: This level is for segmentations that have more than three errors, but these errors are not so severe as to make the segmentation unusable. The errors could be minor misplacements or inaccuracies in labelling. Despite these errors, the segmentation is generally accurate and is considered a pass. | Moderate Quality: Images that show the tongue with adequate quality, but with noticeable imperfections. These may include moderate artifacts, slight motion blur, or some issues with contrast. The overall structure and some of the internal structures of the tongue are discernible, but the image is not ideal for detailed segmentation. (pass+check) |
| **4** | Good Quality: At this level, the overall quality of the segmentation is good, with either minor errors present or perhaps one large error. However, these errors do not substantially impact the overall utility of the segmentation. Pass. | Good Quality: Minor errors in image quality are present in one or more of the domains (CNR, SNR, ringing, etc.). However, these do not significantly impair the visibility of the tongue structure. Pass |
| **5** | Excellent Quality: Segmentation at this level is virtually error-free. It represents the anatomy accurately without any noticeable mistakes. Pass | Excellent Quality: Perfect image quality across all domains, including CNR, SNR, absence of ringing, and other artefacts. The entire tongue is visible, and the image is ideal for segmentation. Pass |

#### Section Three: Intra/Inter rater reliability

Results of the test-retest reliability and inter/intra-rater reliability experiment are shown in Appendix Two Table Three. This experiment was conducted to confirm that the manual segmentation methods (see Appendix 1) could be performed reliably. However, our proposed semi-automatic method does not involve manually drawing entire tongue segmentations.

We calculated Dice overlaps[3], [4] between and within each rater. Dice metrics range from 0, which indicates no spatial overlap between the two sets of manual segmentations, to 1, indicating perfect overlap. As there are no guidelines for cut-offs of Dice overlaps [5] in this context, we estimate that values below 0.5 are weak, values between 0.5–0.6 are satisfactory, values between 0.6–0.7 are considered good, and above 0.7 are excellent.

In general, all raters showed between satisfactory and excellent across the different muscle groups within their own ratings.

Appendix Two, Table Three: Intra-rater reliability

| ***Muscle Labels*** | ***R1 average Dice*** | ***R2 average Dice*** | ***R3 average Dice*** | ***Label Average*** |
|---|---|---|---|---|
| *Genioglossus* | 0.81 | 0.77 | 0.64 | 0.74 |
| *Inferior Longitudinal* | 0.90 | 0.87 | 0.66 | 0.81 |
| *Superior Longitudinal* | 0.88 | 0.90 | 0.64 | 0.81 |
| *Transverse/Vertical* | 0.92 | 0.93 | 0.76 | 0.87 |
| ***Rater average*** | **0.88** | **0.87** | **0.68** | **0.81*** |

*Overall average across rater and labels

Table Three. Intra-rater reliability estimates (using Dice overlaps) for each rater, across two manually labelled datasets, labelled twice, each, per rater.

Appendix Two, Table Four: Inter-rater reliability

| ***Muscle Labels*** | ***R1 average Dice*** | ***R2 average Dice*** | ***R3 average Dice*** | **Label Average** |
|---|---|---|---|---|
| *Genioglossus* | 0.80 | 0.80 | 0.69 | 0.76 |
| Inferior Longitudinal | 0.71 | 0.69 | 0.58 | 0.66 |
| Superior Longitudinal | 0.81 | 0.81 | 0.70 | 0.77 |
| Transverse/Vertical | 0.87 | 0.87 | 0.78 | 0.84 |
| **Rater Average** | **0.80** | **0.79** | **0.69** | **0.76*** |

Table 4. Inter-rater reliability estimates (using Dice overlaps) for each rater, across two manually labelled datasets, labelled twice, each, per rater. Scores were calculated by measuring the average Dice overlap between each pair of corresponding images, between each rater. For example, R1 rated two images, twice: therefore, the

average Dice overlap per muscle reported here represents 12 Dice overlaps between the other raters' images and their own.

#### Section Four: Model selection and IOC covariate results.

To understand the relationships between sex, body measurements (weight and height), and tongue volume (both non-normalised and normalised), we conducted a series of correlations pre- and post-normalisation by IOC. Our goal was to determine the significance of biological variables on tongue volume and the effectiveness of IOC normalisation. A preliminary check for multicollinearity among the predictors—sex (categorical), weight, and height (continuous)—was conducted to ensure the reliability of the regression models. Collinearity was assessed using Variance Inflation Factor (VIF) analysis using the car package in R[59]. Linear regression models were constructed to predict total and normalised tongue volumes based on sex, weight, and height, and to evaluate the role of IOC volume alone on total tongue volume.

Initially, we examined the Variance Inflation Factor (VIF) of our demographic predictors: VIF values: sex (1.54), weight (1.25), and height (1.82) indicated low multicollinearity among all predictors, suggesting that multicollinearity was unlikely to confound the effects of these predictors in subsequent regression analyses (Appendix Two).

Normalised Volume Model: The regression model for normalised volume, including the same predictors, was not significant (Adjusted R-squared: -0.006606, $p = 0.4742$), indicating that sex, weight, and height do not substantially predict normalised tongue volume after accounting for IOC volume.

IOC Model for Comparison: A significant relationship was found between total tongue volume and IOC volume ($F(1,70) = 5.142$, $p = 0.02645$), albeit weaker than the model including demographic variables. This highlights that the size of the oral cavity in volumetric analyses of the tongue is important, but that other biological factors also play a role.

Total Volume Model: The regression model for total volume, which included sex, weight, and height as predictors, was significant (F(3,68) = 8.699, $p < 0.0001$), with sex as a significant predictor (p = 0.0399). This model demonstrates that biological sex is an important determinant of total tongue volume, alongside height which showed a trend ($p$ = 0.1006). We then assessed a regression model with sex as the only predictor of tongue volume. This indicated that female sex significantly reduces total tongue volume by approximately 7696 $mm^3$ compared to male ($p$ = 0.00113).

#### IOC on dataset

IOC measurements across dataset:

We assessed the utility of our IOC technique across datasets, to ensure consistency.

We first examined differences between datasets: **Age:** The ANOVA results for age across different datasets show a significant difference (F(2, 115) = 4.872, p = 0.00931). suggesting age differences between at least two of the datasets. Tukey post-hoc tests revealed a significant age difference between the Sydney and EATT datasets (mean difference = 6.824, p = 0.0107): participants in the Sydney dataset are, on average, older than those in the EATT dataset.

**Weight:** The ANOVA conducted on weight across the datasets did not show a statistically significant difference (F(1, 67) = 2.773, $p$ = 0.101). **Height:** Similarly, no significant differences were found in the heights of participants across the datasets (F(1, 67) = 0.016, $p$ = 0.9). **Sex:** A chi-squared test conducted to examine the distribution of sexes across datasets yielded a significant result ($\chi^2(2) = 6.4518$, $p$ = 0.03972), indicating a significant variation in the proportion of males and females across the datasets. There were more males than females in the EATT and Sydney datasets, while the BeLong dataset presents a more balanced distribution, though still male-dominant.

As reported in Experiment Two, we noticed differences in tongue volume by dataset when uncorrected. Specifically, Sydney vs. BeLong: A significant difference was noted, with

Sydney exhibiting higher mean volumes than BeLong by 1094.17 mm³ (95% CI: 313.40 to 1874.95, $p = 0.003$). Sydney vs. EATT: Similarly, Sydney's mean volumes were significantly greater than those of EATT by roughly 792.94 mm³ (95% CI: 93.72 to 1492.16, $p = 0.022$). EATT vs. BeLong: No significant difference was found between these two datasets (mean difference = 301.23 mm³, 95% CI: -486.63 to 1089.09, $p = 0.641$).

A two-way ANOVA was conducted to assess the impact of muscle type and dataset on tongue volumes, corrected for IOC. The analysis revealed significant main effects for muscle type ($F(3, 456) = 797.26$, $p < 0.0001$) and dataset ($F(2, 456) = 14.78$, $p < 0.0001$), alongside a significant interaction effect ($F(6, 456) = 13.43$, $p < 0.0001$). These results indicate distinct volume differences across muscle types and datasets, with the interaction suggesting the effect of muscle type on volume varies by dataset. This could be related to the demographic details of the dataset, which are significantly different (as above).

We conducted post-hoc Tukey HSD tests to explore these differences. For the transverse/vertical muscles, a significant difference was noted with the Sydney dataset showing reduced volumes compared to BeLong ($p = 0.0083$) and EATT ($p = 0.0039$). The superior longitudinal muscle volumes were higher in the EATT ($p = 0.0004$) and Sydney ($p < 0.0001$) datasets compared to BeLong, with Sydney also showing an increase over EATT ($p = 0.0386$). Genioglossus volumes increased in both EATT ($p = 0.0001$) and Sydney ($p < 0.0001$) compared to BeLong, with no significant difference between Sydney and EATT ($p = 0.4016$). Inferior longitudinal muscle volumes were significantly higher in EATT compared to BeLong ($p < 0.0001$), while Sydney showed a significant decrease compared to EATT ($p = 0.000007$), with no significant difference between Sydney and BeLong ($p = 0.2607$).

Due to a lack of ground truth, we cannot be sure if these discrepancies are due to actual biological variability within the dataset, or differences in scanner hardware, segmentation processing minutiae, or the like. Due to the demographic differences between datasets, it is likely a combination of these factors.

#### Section Five: Tongue position experiment

To test the impact of tongue posture on segmentation validity we scanned one healthy control (F, 31) consecutively, five times, with five different tongue positions. The jaw was always closed, except for position four.

The instructions for the participant were as follows:

1) Rest tongue normally.
2) Depress tip of tongue into the base of the mouth (Bridal).
3) Press the tip and top of tongue to the roof of mouth.
4) Rest entire tongue between teeth, including the back and front of tongue (interdental position).
5) Allow tongue to ‘flop’ to the back of the mouth and retract tongue as far as comfortable (plummy. Cul-de-sac).

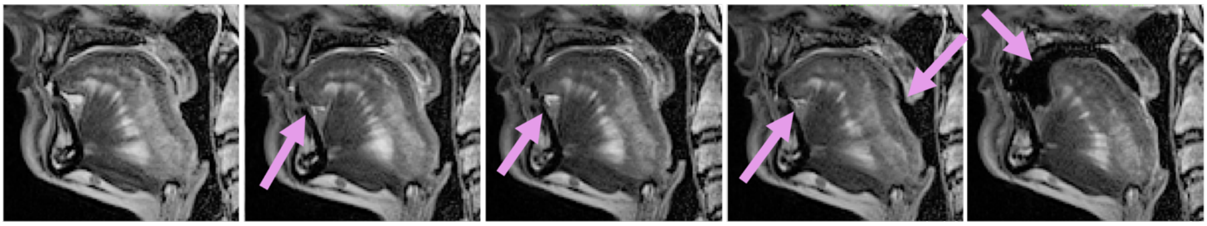

Apeendix Two, Figure 3. sagittal sections of the five tongue movement positions. Pink arrows demarcate areas of changed posture.

We measured the tongue using the semi-automatic method described above, and average percentage changes in muscle volumes between consecutive positions were calculated to determine trends over the positions. A Repeated Measures Analysis of Variance (ANOVA) was conducted to test for statistical differences in muscle volumes across the five positions. The assumption of sphericity was assessed, and a Greenhouse-Geisser correction was applied.
